# Intragenic deletions from whole genome sequencing of 1054 suicide deaths

**DOI:** 10.1101/2025.02.28.25323104

**Authors:** E DiBlasi, AA Shabalin, TJ Nicholas, ET Monson, E Ferris, L Yefimov, S Han, LM Baird, WB Callor, MJ Staley, QS Li, VL Willour, H Coon

## Abstract

Suicide is an urgent public health crisis that claimed over 48,000 lives in the US in 2022. The importance of genetics in suicide risk has been established by classical twin and family studies, and confirmed with recent large genome-wide association studies (GWAS). While the GWAS are beginning to reveal genetic risk due to common variants each with small effect on liability, these results explain only a fraction of the genetic risk. As with other complex health conditions, some of this unexplained risk is likely due to rarer variants with larger effect on liability. Using whole genome sequencing (WGS) data from 1,054 population-ascertained Utah suicide deaths, we investigated intragenic deletions as a class of genomic variation highly likely to disrupt gene function. To minimize the chance of false positive results, studied deletions were limited to those found in three large publicly-available control datasets (1000 Genomes, GnomAD, and Centers for Common Disease Genomics). Additional internal replication also required deletions to occur at least twice in WGS from an initial cohort of 670 suicide deaths then again in a second cohort of 384 suicide deaths. All results meeting these filters were manually validated. There were 11 validated deletions with at least 2-fold increase in frequency over occurrence in controls (range 2.28 to 4.46). These results implicated genes associated with risk of mental health conditions (*MPST, IL4R, CDH13*), epilepsy (*CLCA4*), intellectual disability (*ZNF44*), neuronal function (*OSBPL2*), metabolic function (*FBOX36*), lipid metabolism (*TM9SF3*), immune related functions (*PIPOX, IL4R*), and transcriptional repression (*ZHX3*). SNPs in genes implicated by the deletions have also been associated with mental health conditions, neuronal function, immune response, and other critical biological pathways including neuroinflammation and cellular response to stress. Demographic and clinical associations of suicide deaths with specific genetic deletions, highlight the prevalence of mood, anxiety and bipolar disorders and variations in age at suicide death among affected individuals. This work is the largest genome-wide analyses of WGS variation in suicide deaths to date. Pending replication, results will guide future functional studies with the eventual goals of increased understanding of mechanisms leading to risk.

## Introduction

In the U.S., the suicide death rate is the highest it has been in over 80 years (Curtin et al., 2023); suicide prediction and prevention are top public health priorities. Identification of risk factors leading to suicide death remains challenging due to the complexity of suicide risks; however, genetic factors comprise one important aspect of risk that may allow substantial progress in bridging this knowledge gap, with estimates of heritability of approximately 50% (McGuffin et al., 2001; N. L. Pedersen & Fiske, 2010; Statham et al., 1998).

Recent genome-wide results based on large samples have begun to reveal common genetic factors associated with suicide attempt and mortality (Docherty et al., 2023; Kimbrel et al., 2023; Q. S. Li et al., 2022; Mullins et al., 2022). Biological pathway analyses among GWAS with significant loci implicate nervous system function and development as well as immune function, neuroinflammation and cellular response to stress. While these findings represent significant progress, they also highlight the complex polygenic nature of suicidal behavior and the need to detect additional risk loci. GWAS studies provide estimates of SNP heritability ranging from 3.9 – 5.7%, considerably less than the heritability of 50% from the classical behavioral genetics designs involving aggregated family and twin studies. This “missing heritability” likely derives from effects of rare genomic variation and from the contribution of gene x gene effects (Génin, 2020).

While extensive research has focused on common genetic variants in psychiatric phenotypes, far less attention has been given to rarer genetic variation, particularly structural variation (Owen & Williams, 2021). Structural variants (SVs) are a broad class of genome variation that is extremely diverse in type and size (typically 50 bp to several Mb). SVs includes copy number variants (CNVs), such as deletions and duplications, simple re-arrangements (e.g., inversions), mobile element insertions and other complex rearrangements (multiple combinations of SVs) (Ho et al., 2020). SVs are present in all human genomes often have severe functional consequences due to their ability to disrupt genes, cause gene fusions, rearrange regulatory elements and alter gene dosage (Weischenfeldt et al., 2013). SVs have historically been harder to detect than common variation data due size and limitations in associated sequencing technologies and detection algorithms (Ho et al., 2020). Thus, like rarer SNVs, structural variants have largely been excluded from genotyping arrays used in large-scale genetic studies. This gap in knowledge is significant, as other types of genetic variation may play a crucial role in understanding the genetic underpinnings of complex psychiatric conditions (Andreassen et al., 2023). Indeed, studies have begun to implicate rare variation associated with risk of suicide death (DiBlasi et al., 2021; DiBlasi et al., 2021). However, it is crucial to note that these results await replication in larger, independent cohorts. Further research is needed to validate and expand upon these initial discoveries, potentially opening new avenues for suicide prevention and targeted interventions.

The current study provides results from a genome-wide screen of whole genome sequence (WGS) data in a large (N=1,054), population-ascertained sample of suicide deaths. These aspects address previous gaps in studies of rare variation, including small sample sizes, reliance on genotyping rather than WGS data, and/or ascertainment within individuals with specific clinical diagnoses. Using WGS data, emerging approaches can be used to more reliably detect and call SVs (Ho et al., 2020). The current genome-wide investigation focuses on deletions (a class of SVs) overlapping coding sequence of genes; these represent genomic events most likely to affect gene function. We have chosen to focus on deletions as the category of SVs with the most robust ascertainment as they are more amenable to detection by available robust computational tools. We use an independent internal replication strategy for deletion results in our sample, and present cross-associations implicated by previous rare variant studies and by current large GWAS of a range of suicide outcomes.

## Methods

### Samples and data linking

Samples from Utah suicide deaths used in this study were obtained through a long-term collaboration with the Utah State Office of the Medical Examiner (OME). Suicide determination was made by the OME following detailed investigation of the scene and circumstances of death. High quality DNA was extracted from whole blood as previously described (Coon et al., 2020). Identifiers from suicide deaths were securely transferred by the OME to staff at the Utah Population Database (UPDB, https://uofuhealth.utah.edu/huntsman/utah-population-database). The UPDB is a statewide database with >27 million data records, including demographics, electronic health records data, and genealogical records. After linking suicide deaths to health data, identifiers were stripped by UPDB staff prior to secure data transfer to the research team. This study was approved by Institutional Review Boards at the University of Utah, Intermountain Healthcare, and the Utah Department of Health and Human Services.

The 1,054 suicide deaths in this study were selected for whole genome sequencing (WGS) based on factors associated with increased likelihood of greater genetic risk, including younger age at death and evidence for significant extended familial risk of suicide death using Utah genealogical data (Coon et al., 2022), and also diagnoses of bipolar disorder, which is associated with high risk of suicide death (Miller & Black, 2020). Processing of WGS data was done in two waves, the first including 674 Utah suicide deaths and the second including 384 additional Utah suicide deaths.

### Control data

Joint processing of WGS data from Utah suicide deaths was done with individual WGS data from 1,241 controls chosen to most closely match the predominantly Northern European ancestry of the Utah population from three sources: 1) 622 were from the 1000 Genomes Project (1000G) (Byrska-Bishop et al., 2022)2) 512 were unrelated individuals from the Centre d’Etude du Polymorphisme Humain (CEPH) genetic reference families (Dausset et al., 1990); and 3) 96 were from a Utah study of longevity (Tschanz et al., 2004).

### Relatedness and ancestry

Pairwise relatedness was tested using PLINK (Purcell et al., 2007). Among suicide deaths, sample pairs with third-degree relatedness or closer were retained to allow for correction of initial results and prioritization for potential future analyses if familial transmitted variants were observed. Among controls, only one randomly selected case for each pair with third degree or closer relatedness was retained for analysis. Any controls related to suicide deaths at the level of third degree or closer were eliminated.

Ancestry was determined through comparison of genotypes to reference population data available in the 1000G. Small numbers of non-European samples were expected in the suicide death cohort, as samples were ascertained from the Utah population which is predominantly White. In addition, selection of suicide deaths for WGS using increased extended familial risk was expected to further elevate the percentage of suicides with European ancestry, as familial risk assessments required the use of UPDB genealogical data which derives primarily from family founders of European descent. In this study, all suicide deaths were retained regardless of ancestry. Checks were then implemented to determine the extent to which any result was driven by non-European carriers of the deletions.

### Whole genome sequencing pipeline

WGS data was generated on the 1,054 Utah suicide deaths and 1,241 jointly called control samples by using Illumina NGS technology with an average read depth of ∼30x. Alignment and variant calling and joint genotyping of suicide deaths and control WGS datasets was performed at the Utah Center for Genetic Discovery (UCGD) Core Facility, part of the Health Sciences Center Cores at University of Utah. The UCGD pipeline (v 2.13.77) called variants using the Sentieon software package (Freed et al., 2017), which incorporates GATK best practices (Depristo et al., 2011; Van der Auwera et al., 2013) with other innovative elements. Use of this UCGD pipeline has been described in detail elsewhere (Nicholas et al., 2022). Briefly, sequence reads were aligned to GRCh38 (Genome Reference Consortium Human Build 38) using BWA-MEM (H. Li & Durbin, 2009). The Haplotyper algorithm in Sentieon (Freed et al., 2017)was used to produce genomic Variant Call Format (gVCF) files. Suicide death gVCF files were then combined and jointly processed with data from the 1,241 individual control samples. Processing and analysis of data was done in two waves, one with 670 suicide deaths and the second with 384 suicide deaths. However, the final VCF file with suicide deaths and controls was recalled across both waves of suicide data together with controls to limit false positive calls.

### Structural variant (SV) calling

Detection, attribution, and filtering of structural variants (SVs) was done using Smoove (version 0.2.6; https://github.com/brentp/smoove) and Manta (version 1.6.0) (Chen et al., 2016), again using the GRCh38 reference genome. Smoove produces a VCF file that contains deletions, duplications, inversions, and unclassified breakends (BNDs). Overlapping genes (Ensembl GRCh38.p12) and Duphold (B. S. Pedersen & Quinlan, 2019) quality metrics were added to the Smoove-generated VCF files. SVAFotate (https://github.com/fakedrtom/SVAFotate) was also used to add population SV allele frequencies from control datasets for comparison. SV frequencies from WGS datasets were available from the large publicly available cohorts described above (CCDG, GnomAD, 1000G, and CEPH, N=603). The reciprocal overlap fraction requirement of 0.8 was used to match control datasets in SVAFotate. This study focuses on the deletions, as these are the most robustly detected class of SV.

### Additional filtering criteria for deletions

Additional filtering was performed using Slivar (B. S. Pedersen et al., 2021) to minimize artifacts and maximize potential functional deletions. Criteria included: 1) removing deletions <100 base pairs (bp) and >1 megabase (Mb) as likely artifacts; 2) requiring deletions to overlap an exon using Smoove_gene INFO values (https://github.com/brentp/smoove); 3) requiring deletions to be present in at least two suicide deaths; and 4) requiring deletions to pass duphold (B. S. Pedersen & Quinlan, 2019) metrics (Duphold_pass flag and fold change DHFFC <0.7). Estimation of population frequencies of deletions were made using four large public datasets: 1) the Genome Aggregation Database comprising a subset of individuals from studies of adult-onset common diseases (Collins et al., 2020); 2) Centers for Common Disease Genomics (CCDG) comprising 10,738 individuals from studies of cardiovascular, neuropsychiatric, and immune-mediated diseases (Abel et al., 2020); 3) 1000G comprising 3,202 individuals (Byrska-Bishop et al., 2022); and 4) CEPH comprising 603 unrelated individuals in 34 multigenerational families (Dausset et al., 1990). Deletions were required to have frequency of <0.2 in each of these large control datasets. These criteria were required to be independently met in the first set of WGS data from 670 Utah suicides, and then also in the second set of 384 Utah suicide WGS data. This internal replication was followed to increase the robustness of our design. Frequencies of deletions were compared to observed frequencies in the jointly processed controls using exact binomial tests.

### Manual validation of deletions

Smaller deletions (less than 5,000 bp) were verified by PCR and subsequent electrophoresis of the PCR product on agarose gels. First, primers flanking the putative deletions were designed using the Primer3 program (v. 0.4.0). 50 ng of template DNA was then amplified with these primers in a PCR reaction using Q5®High-Fidelity DNA Polymerase (New England Biolabs®) per manufacturer’s recommended conditions. PCR product was subsequently elecrophoresed on 1XTBE 2% agarose gels in the presence of Ethidium Bromide and then visualized on a Fotodyne™ FOTO/Analyst™ Workstation. The deletion was verified when product bands of two different sizes were present after PCR, one band being the larger wild type allele and the other band being a smaller deleted allele. Larger deletions (greater than 5,000 bp) were verified using pre-designed Taqman™ Copy Number Assays purchased from Thermo Fisher Scientific. Quadruplicate reactions were carried out per manufacturer’s recommendations, and the data was analyzed with CopyCaller® Software.

Additional validation was done using SAMPLOT (Belyeu et al., 2021) to visualize the deletions *in silico* for comparison with manual validation results and to obtain indications of read depth sequence alignments in the region of the deletion. SAMPLOT images were reviewed for call and confidence intervals. Specifically, each deletion was inspected in the called region to have a loss in the depth of coverage and split or discordant read signatures.

### Characterization of SNPs within deletions and of implicated genes

Potential functional impact of SNPs within each significant deletion was characterized using public data from the GTEx consortium (https://gtexportal.org/). Additional characterizations of the genes implicated by the deletions included publicly-available genetic associations from the GWAS Catalog (Sollis et al., 2023), and drug interaction data (Cannon et al., 2024; Gaulton et al., 2012).

### Clinical data

Once SVs were identified and validated, additional electronic health records (EHR) data linked to suicide deaths were then used to assess for clinical associations of deletions with co-occurring psychiatric and physical health conditions among carriers of prioritized variants. Diagnostic data from the International Classification of Diseases (ICD-9 and ICD-10) system in the EHR was tabulated for suicides with deletions. Diagnoses were collapsed into phecodes (Denny et al., 2010), curated groups of ICD codes created to capture clinically meaningful phenotypes, using the hierarchical classification of diagnoses in the PheWAS catalog (PheCode Map X) (Shuey et al., 2023), Diagnoses that occurred within one week prior to death were excluded to eliminate diagnoses associated with the final fatal suicide event rather than those reflecting prior risk. Diagnoses relating to “Mood [affective] disorders” (phecode MB_286); “Bipolar disorder” (phecode MB_286.1); “Psychoactive substance related disorders” (phecode MB_280); “Pain” (phecode SS_809); “Psychotic disorder” (phecode MB_287); “Anxiety and anxiety disorders” (phecode MB_288) and “Sleep disorders” (NS_333) were studied.

## Results

### Sequencing quality control

Overall, WGS in the Utah suicide deaths was of high quality, with coverage from 25x to 48x across both batches, averaging 30x. In the first set of 674 suicides, four were omitted due to low quality, leaving 670 for analysis. In addition, one sample in this batch had relatively low alignment (77.6%), four had alignment between 80-90%, and four had alignment between 90-95%. All of these were retained but scrutinized at the manual validation stages. In the second set of 384 suicides, one suicide selected for sequencing failed at the library prep phase, and was replaced with another suicide death sample, One sample in this second set had relatively low alignment (74.2%), two others had alignment between 80-90%, and three others had alignment between 90-95%. All of these were again retained for analysis and scrutinized at the manual validation stages.

For two of the suicides in this first WGS set and three in the second set, sex in the data records did not match genetic sex. Because all of the samples collected in our protocol are suicide deaths, the five samples with sex mismatches still represent suicides, so they were retained for tests of genetic ancestry and deletion frequency analyses. However, they were omitted for other phenotypic analyses. The suicide death cohort size was therefore 1,054 for frequency tests, but 1,049 for other phenotypic analyses.

### Selection characteristics of suicides for WGS

Descriptive data of the suicides with WGS are shown in Table 1. A study design decision to further enhance our selection for bipolar disorder between cohorts 1 and 2 resulted in inclusion in the second cohort of more individuals with older age at death and also more females. Tests between the initial cohort of 670 and the second cohort of 384 suicides resulted in significant differences in these demographic characteristics (age test: t=10.93, p<0.0001; sex test: chi-square=11.75, p=0.0006). Our overall WGS sample selection strategy also resulted in the combined cohort of Utah WGS suicides being significantly younger at death than in 5,386 other Utah suicides with genotyping but without WGS (32.13 years vs. 42.26 years; p<0.0001). The full sample of suicides with WGS were also significantly more female than in the larger genotyped cohort (27.84% female vs. 19.97% female; p<0.0001).

**Table 1.** Demographic characteristics of Utah suicide deaths with WGS data.

| Cohort | N with WGS | % female | Avg. age at death in years (sd) | % European ancestry | N with bipolar disorder |
| --- | --- | --- | --- | --- | --- |
| Cohort 1 | 670 | 24.25% | 28.87 (sd=13.08) | 97.72% | 145 |
| Cohort 2 | 384 | 34.12% | 37.84 (sd=12.21) | 98.58% | 180 |
| All suicides with WGS | 1,054 | 27.84% | 32.13 (sd=13.47) | 98.10% | 325 |

Note. Demographic age and sex data and clinical data omits 5 suicides known to have mismatched sex between records and genotyping. As all collected samples were suicide deaths, these 5 cases were included for ancestry tests.

Utah suicides with WGS were of predominantly European ancestry (98.10%). There were 15 WGS suicide deaths with <90% EUR ancestry (between 51% and 84%; see Supplemental Table 1). These were retained in our initial analyses, with sensitivity tests to determine the extent to which any results were driven by occurrence of variants in non-European case samples. Control samples were chosen to match expected primarily European ancestry of population-ascertained cases.

Relatedness tests among the suicide deaths indicated nine third-degree relative pairs (cousins), five second-degree pairs (grandparent-grandchild or avuncular or half-siblings), and six first-degree relative pairs (siblings or parent-child). All related suicides were retained. Our analysis design dictated that if both relatives of a relative pair were found to share a variant or deletion, statistics for the variant/deletion would be re-adjusted, and the variant/deletion considered for potential future familial analyses.

Control data was also checked for relatedness. Among controls, there were two third-degree relative pairs, one second-degree pair, and two first-degree pairs. One random related control from each relative pair was deleted for the analysis. Finally, there were five suicide deaths where controls were found with a third degree relationships, and two suicides where controls were found with second degree relationships. These related controls were not considered in the analyses.

### SV discovery and filtering

In the initial set of 670 suicide deaths, used as the discovery set, 58,012 deletions were discovered. While additional structural variants were apparent (e.g., insertions, inversions), deletions are the most common event and also most robustly detected, so further analyses focused on this class of structural variant. Of the 58,012 deletions, 7,616 crossed an exon using the Smoove_gene info prediction. Our next filters requiring the Duphold pass flag, fold change DHFFC<0.7, and presence in at least two suicide deaths resulted in 831 deletions. Using the SVAFotate tool, each deletion was compared to the public data cohorts (GnomAD, CCDG, 1000G, and CEPH). Deletions were deemed to be present in controls if there was at least 0.8 overlap in deletion boundaries. We only retained those with a frequency of <20% across these control cohorts (to eliminate common events), resulting in 391 deletions. Of these 391, 29 were >1Mb, and 42 were <100bp; these were omitted as likely artifacts.

After all of these filtering steps, there were 320 deletions, 113 of which were not found in any of the control cohorts; most of these unique deletions (N=96) were called as imprecise. We retained only the 207 found in the large control datasets. Our next step was to match these deletions to the second set of 384 suicide deaths. We required precise base-pair matching of the start and stop of the deletion in two suicides in the second set of 384 suicides for this replication step. Of the 25 deletions that met this requirement, there were 14 deletions with at least two occurrences in this second set, and with at least a two-fold increase in frequency of each deletion over and above its frequency in jointly processed controls. Final determination of significance was made only after manual validation of each deletion in each case, as detailed below. Eleven deletions were significant after manual validation (Table 1).

### Manual validation of deletions

Validation for seven of the smaller deletions was done using PCR (*PIPOX, TM9SF3, ZNF44, OSBPL2, ZHX3, APOOL, MPST*; see Supplemental Tables S1-S2 for primers and details). Primers for the *LILRA1* deletion (chr19:54601316-54601631) were not possible to define given extensive homology across the region of the deletion with three other regions on chromosome 19. For the *APOOL* deletion (chrX:85087992-85088221), only seven of the 16 suicides clearly showed the deletion. These two deletions were therefore not studied further in this analysis. Validation of *MPST* showed evidence of varying copy numbers of this SV in both suicides and controls. Though results still indicate case-control differences (see Supplemental Figure S1), this variation indicates interpretation of this deletion should be approached with caution. For the other deletions validated by PCR, all suicides determined to have the deletion by silico methods (Samplot; Belyeu et al., 2021) showed validation with PCR with the exception of one sample for the *TM9SF3* deletion, leaving 10 suicides with that deletion (case frequency = 0.00476, fold increase over controls = 3.33).

For the larger deletions requiring Taqman validation, *FBXO36, LMNTD1, CLCA4, FBX036,* and *IL4R* were manually validated for all suicides with these deletions. For *CDH13*, two of the 20 suicides initially identified as having the deletion failed to validate, leaving 18 with the deletion (case frequency = 0.000857, fold increase over controls = 2.06).

Table 2 gives details of each validated deletion. These 11 deletions were significantly elevated over the frequency in jointly-processed WGS data from controls at a false discovery rate (FDR) of 0.05. Frequencies from the publicly-available GnomAD “non-neuro” subset (Gudmundsson et al., 2021), which omit genomes from individuals with neuropsychiatric conditions, are also shown for comparison.

**Table 2.**
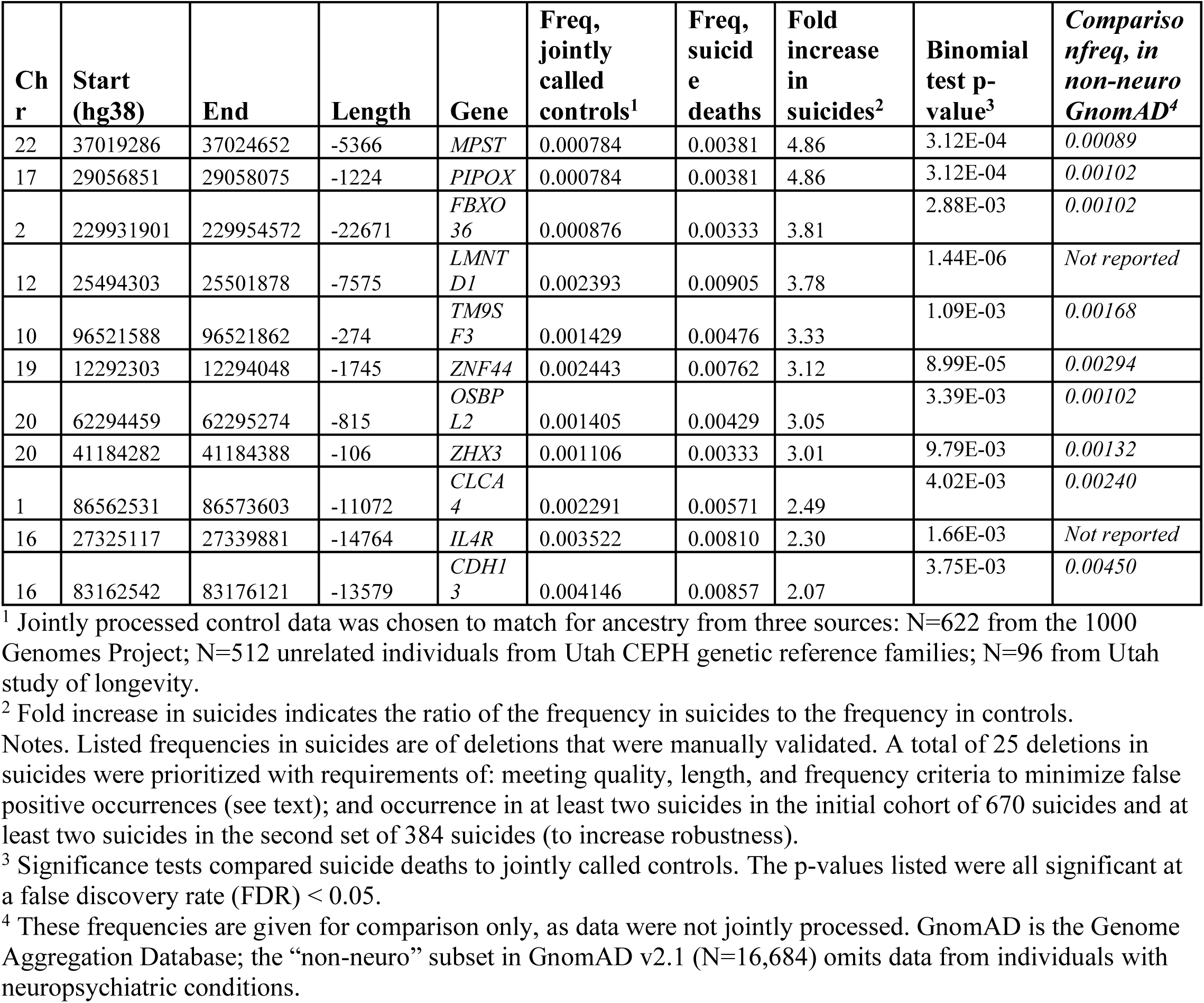
Manually validated deletions present in at least two suicides in each of the two suicide death cohorts and showing > two-fold increase in frequency over jointly processed control sequence data.

### SNP-level and gene-level associations of significant deletions

For the 11 significant deletions, tissue associations are summarized in Table 3 for SNPs within each significant deletion with overlapping in GTEx data (https://gtexportal.org/home/). Detail of the GTEx SNP-level data is given in Supplemental Table S3. At the gene level, Table 3 gives a summary of associations for each gene implicated by the 11 deletions listed in the GWAS catalog (https://www.ebi.ac.uk/gwas/home), and gene-drug interactions from data in the Drug-Gene Interaction Database (DGIdb; https://www.dgidb.org/).

**Table 3.** Additional characterizations of implicated genes.

| Gene implicated by deletion | GTEx expression of SNPs within deletion regions<sup>1</sup> | GWAS Catalog associations | DGIdb gene-drug interactions; DRUGBANK drug targets |
| --- | --- | --- | --- |
| *MPST* | No del SNPs overlap with GTEx | Hemoglobin, gut microbiome, metabolite levels, putamen | DGIdb: Thyrozine (thyroid hormone levels) |
| *PIPOX* | 4 SNPs, adrenal gland expression | MDD, Depression, AD, BMI, accelerated aging, language, smoking heaviness, smoking initiation | DRUGBANK DB00145, Glycine (nutraceutical; used for stroke, MI; fast inhibitory neurotransmitter, possible antipsychotic activity) |
| *FBXO36* | 19 SNPs, 4 with brain expression | Triglycerides, creatinine, fat mass, BMI | None |
| *LMNTD1* | 4 SNPs, testis expression | Gut microbiome, monocyte count | DGIdb: Capecitabine (cytotoxic agent, cancer treatment) |
| *TM9SF3* | No del SNPs overlap with GTEx | Bone density | None |
| *ZNF44* | 4 SNPs, 3 expressed in brain | Alcoholic liver disease | None |
| *OSBPL2* | 1 SNP expressed in testis | Bipolar, platelet volume, nicotine dependence, reticulocyte fraction, cortical folding, educational attainment | Non |
| *ZHX3* | No del SNPs overlap with GTEx | Unipolar depression, daytime napping, sleep duration, MDD, depressive symptoms, drinks per week, migraine, triglycerides, cholesterol, CAD, BMI, waist-hip ratio | None |
| *CLCA4* | 11 SNPs, all associated with brain expression | IgG glycosylation, macrophage inflammatory protein 1b levels | None |
| *IL4R* | 27 SNPs, 17 with brain expression | Asthma, allergy, C-reactive protein, respiratory diseases, IgE levels, eosinophil counts | DGIdb: Sunitinib (kinase inhibitor); DGIdb and DRUGBANK (DB12159): Dupilumab (monoclonal antibody, asthma, COPD, dermatitis); DRUGBANK: DB05078m AER001 (asthma treatment) |
| *CDH13* | No del SNPs overlap with GTEx | ADHD, smoking initiation, clozapine response, amphetamine response, SZ, AD onset age, educational attainment, cognitive ability, CAD, blood pressure, adiponectin levels, COPD treatment response, BMI, antibody response to COVID-19 vaccine | Deutetrabenain (tardive dyskinesia/chorea in HD, NOTE: may increase suicidality); Dextro-amphetamine sulfate (ADHD, cognitive dysfunction) Clopidogrel (plavix; anti-thrombotic) |

<sup>1</sup> Further detail of SNP names, p-values, and GTEx-associated tissues appears in Supplemental Table S3. See Supplemental information for GWAS Catalog references.

### Phenotypic associations

There were no related pairs among the suicide deaths sharing validated deletions in Table 2, nor were any of these cases among those with known sex mismatching. None of the validated deletions occurred in samples from individuals with <95% European ancestry with the exception of the deletion in *TM9SF3* on chromosome 10. This deletion was carried in two suicides with <95% European ancestry, one with 84.1% European ancestry, 15.0% Admixed American ancestry, and 0.9% South Asian ancestry, and the other with 81.3% European ancestry, 17.9% East Asian ancestry and 0.8% Admixed American ancestry. Removing these two occurrences from the data lowered the observed frequency for this frequency to 0.00381, leaving a fold increase of 2.67 and a binomial test significance of 1.19E-02 which remained significant at FDR (0.05).

Table 4 presents demographic and clinical associations of suicides carrying each deletion compared to overall frequencies in all suicide deaths with WGS data. Results are aggregated to protect confidentiality. 1,030/1,053 samples with WGS data had ICD diagnoses and could be assessed for phecode phenotypes. Of the samples with WGS and ICD data, mood disorders diagnoses were prevalent in 69.1% of suicide deaths, anxiety disorder (50.5%), bipolar disorder (42%), and substance related disorders (48.2%) were also common diagnoses. All suicide deaths with identified deletions in *TM9SF3* and *ZHX3* had mood disorder diagnoses. Suicide deaths with a deletion in MPST had the youngest average age at death at 28.3 years.

**Table 4.** Demographic and clinical characteristics of suicide deaths with validated deletions.

| All samples and by gene implicated by deletion | Avg. Age at death (SD) | % Female | % Anxiety and anxiety disorders | % Bipolar disorder | % Mood [affective] disorders | % Pain | % Psychoactive substance related disorders | % Psychotic disorder | % Sleep disorders |
| --- | --- | --- | --- | --- | --- | --- | --- | --- | --- |
| *MPST* | 28.3 (11.8) | 37.5 | 44.4 | 22.2 | 66.7 | 11.1 | 33.3 | 11.1 | 11.1 |
| *PIPOX* | 36.1 (16.7) | 12.5 | 62.5 | 37.5 | 62.5 | 50.0 | 37.5 | 12.5 | 50.0 |
| *FBXO36* | 28.3 (15.0) | 14.2 | 28.6 | 14.3 | 42.9 | 57.1 | 28.6 | 0 | 28.6 |
| *LMNTD1* | 35.8 (13.1) | 36.8 | 57.9 | 47.4 | 73.7 | 36.8 | 42.1 | 21.1 | 21.1 |
| *TM9SF3* | 32.7 (11.5) | 40.0 | 55.6 | 55.6 | 100 | 55.6 | 55.6 | 22.2 | 22.2 |
| *ZNF44* | 37.1 (13.6) | 43.8 | 68.8 | 31.3 | 87.5 | 31.3 | 56.3 | 37.5 | 50.0 |
| *OSBPL2* | 36.3 (17.4) | 33.3 | 44.4 | 66.7 | 66.7 | 33.3 | 44.4 | 33.3 | 55.6 |
| *ZHX3* | 39.00 (10.4) | 42.9 | 71.4 | 71.4 | 100 | 42.9 | 71.4 | 28.6 | 14.3 |
| *CLCA4* | 37.7 (13.0) | 16.7 | 66.7 | 85.7 | 83.3 | 50.0 | 50.0 | 41.7 | 66.7 |
| *IL4R* | 38.1 (13.5) | 23.5 | 53.0 | 41.2 | 76.5 | 41.2 | 58.8 | 29.4 | 29.4 |
| *CDH13* | 32.8 (13.1) | 22.2 | 44.4 | 55.6 | 66.7 | 61.1 | 44.4 | 16.7 | 27.8 |
| All suicides with WGS | 32.1 (13.5) | 27.8 | 50.6 | 42.0 | 69.1 | 36.02 | 48.2 | 21.3 | 30.1 |

## Discussion

This study represents the most extensive genome-wide investigation of structural variation, specifically intragenic deletions, in suicide decedents to date. Our analysis of whole genome sequencing (WGS) data from 1,054 suicide cases and 1,241 controls identified 11 validated deletions with significant enrichment in suicide deaths compared to controls. The use of WGS data allowed for a comprehensive analysis of deletions, overcoming limitations of previous studies that relied on genotyping arrays which do not contain this genetic information. We also employed an internal replication strategy and manual validation of deletions. These deletions are predicted to impact genes associated with mental health conditions, neuronal function, immune response, and other critical biological pathways, offering new insights into the genetic underpinnings of suicide risk.

Results highlight the potential role of rare intragenic deletions in suicide risk, a dimension less explored compared to common single nucleotide polymorphisms (SNPs). The identified deletions overlap genes involved in diverse biological functions, such as mental health (*MPST, IL4R, PIPOX, CDH13*) (Bonk et al., 2024; Ruggeri et al., 2015; Salatino-Oliveira et al., 2015; Schoormans et al., 2012), neuronal function (*OSBPL2*), and immune related functions (*PIPOX, IL4R*) (Paulovičová et al., 2021; Vogelaar et al., 2018). Notably, several of these genes, such as *PIPOX*, *ZHX3, CDH13* (Børglum et al., 2014; Coleman et al., 2020; Fabbri et al., 2019; Mullins et al., 2021) have been implicated in previous GWAS related to psychiatric and neurodevelopmental disorders.

The association of deletions with *CLCA4* (He et al., 2021), linked to epilepsy, and *ZNF44* (Bassuk et al., 2013), associated with intellectual disabilities, also highlights the potential overlap between suicide risk and other neurodevelopmental and neurological conditions. Additionally, metabolic function (*FBXO36*) (Riveros-Mckay et al., 2020), lipid metabolism (*TM9SF3*) (Zhu et al., 2022), immune related functions (*PIPOX, IL4R*), and transcriptional repression (*ZHX3*) (Igata et al., 2022). *LMNTD1* is involved in structural molecule and may be associated with cancer risk and motor memory (Nguyen et al., 2022).

### Study Limitations and Future Directions

Most of our suicide death cohort is of European ancestry, which may limit the generalizability of the results in other populations. Since the observed deletions were found predominantly in individuals of European ancestry, further studies in diverse populations are necessary to validate these findings across different genetic backgrounds. Replication in larger and independent cohorts will be needed to confirm these findings. Additionally, while our study provides support for the role of intragenic deletions in suicide risk, characterizing the biological impact of these deletions on gene expression and function, and exploring potential interactions with other genetic and environmental risk factors.

The next steps in this work will include a broader investigation that includes non-coding deletions, other structural variant types (e.g., duplications, inversions), and a more comprehensive assessment of additional putatively functional single nucleotide variants (SNVs). Future work will also include interactions of common polygenic backgrounds and/or environmental exposures with the occurrence of rare variants. The study of rare genetic variation leading to suicide risk will increase understanding of biological mechanisms and serve to identify potential drug targets with the ultimate goal of improved prevention and personalized interventions.

It is important to emphasize that having a deletion associated with suicide death does not deterministically confer suicide risk but rather represents just one component of multifaceted genetic and environmental interactions. Complex traits such as suicide are influenced by multiple genetic variants, each often exerting small, additive, or interactive effects, along with epigenetic modifications and environmental factors that modulate gene expression and phenotype manifestation. A single deletion may increase risk, but its impact is contingent on broader genomic context, gene-gene interactions, and external influences. Thus, while discovering deletions provides valuable insight into disease mechanisms, it should be interpreted as one piece of a larger, intricate puzzle rather than a definitive predictor of the trait

## Conclusion

This work advances our understanding of the genetic factors contributing to suicide risk by examining the role of rare intragenic deletions. Our findings highlight the importance of integrating structural variant analysis with existing knowledge from GWAS and other genetic studies. As the field progresses, further research and replication in independent cohorts will be essential to confirm these results and understand the underlying biological mechanisms, ultimately contributing to improved strategies for suicide prevention.

## Supporting information

Supplemental Informaton

samplots

validation images

## Data Availability

All data produced in the present study are available upon reasonable request to the authors

## Acknowledgements

This work was supported by the National Institute of Health (HC, grant number R01MH122412, R01MH123489; AD, grant number R01MH123619; AVB, grant number R01ES032028); the American Foundation for Suicide Prevention (ED), the Brain & Behavior Research Foundation--NARSAD (ED, grant number 28132; AS grant number 28686); and the Clark Tanner Foundation (HC, AS, EM, AVB). Partial support for all datasets housed within the Utah Population Data Base is provided by the Huntsman Cancer Institute (HCI), http://www.huntsmancancer.org/, and the HCI Cancer Center Support grant, P30CA42014 from the National Cancer Institute. Research was supported by NCRR grant “Sharing statewide health data for genetic research” R01RR021746 with additional support from the Utah Department of Health and Human Services and the University of Utah. We thank University of Utah Health Data Science Services for data and analytics support, and the University of Utah Pedigree and Population Resource and the University of Utah Health Enterprise Data Warehouse for establishing the Master Subject Index between the Utah Population Database and the University of Utah Health Sciences Center. DNA extraction was performed by the University of Utah Center for Clinical and Translational Science supported by the National Center for Advancing Translational Sciences of the NIH (grant number UL1TR002538). Genotyping was performed by the University of Utah Genomic Core (UL1TR002538) and by Illumina, Inc. with support from Janssen Research & Development, LLC. The support and resources from the Center for High Performance Computing at the University of Utah are gratefully acknowledged.

