## Supplemental Tables for "Intragenic deletions from whole genome sequencing of 1054 suicide deaths"

**Supplemental Table S1. Primers for manual validation of shorter deletions.**

| chr | gene | deletion | length | forward primer | reverse primer |
|---|---|---|---|---|---|
| 20 | *ZHX3* | 41184282-41184388 | 106 | CCAAACCATAGGAGGCTCT | GGATGCTTTACCATTCTCTTTAGTT |
| X | *APOOL* | 85087992-85088221 | 229 | AACACACAAACTGTGAATGCAG | GAGGGAAACACACCTTTCCA |
| 10 | *TM9SF3* | 96521588-96521862 | 274 | TGCATTGTAGAGAGTACACTTCT | AGACCAGCTCTCTTCAGTGA |
| 19 | *LILRA1* | 54601316-54601631 | 315 | not possible (dropped) | not possible (dropped) |
| 20 | *OSBPL2* | 62294459-62295274 | 815 | GCCAGGGATTGCCTTCTAA | GGCCTGAGTAACCTGGACTC |
| 17 | *PIPOX* | 29056851-29058075 | 1224 | GAAGAGGGCACAAGAACTGG | AAAAAGCAGACTTGGCATGG |
| 19 | *ZNF44* | 12292303-12294048 | 1745 | GTGGGCATTCCCAGCTACT | GCTTCTCCCCAGATTGTGC |
| 22 | *MPST* | 37019286-37024652 | 5366 | TTGGGCTTAGAATCCCCTTT | CCAGGTTCTCCTTGATGTCC |

**Supplemental Table S2. Quality metrics and manual validation results**.

| Chr | POS | end | length | gene | Quality Score | strands | Manual validation |
|---|---|---|---|---|---|---|---|
| 22 | 37019286 | 37024652 | -5366 | MPST | 761.5 | STRANDS=+-:123 | Validated with variability |
| 17 | 29056851 | 29058075 | -1224 | PIPOX | 230.7 | STRANDS=+-:18 | Validated |
| 19 | 54601316 | 54601631 | -315 | LILRA1 | 548.6 | STRANDS=+-:12 | Unable to validate |
| 2 | 229931901 | 229954572 | -22671 | FBXO36 | 395.6 | STRANDS=+-:14 | Validated |
| 12 | 25494303 | 25501878 | -7575 | LMNTD1 | 1070 | STRANDS=+-:252 | Validated |
| 10 | 96521588 | 96521862 | -274 | TM9SF3 | 755 | STRANDS=+-:96 | 10/11 Validated |
| 19 | 12292303 | 12294048 | -1745 | ZNF44 | 749.6 | STRANDS=+-:172 | Validated |
| 20 | 62294459 | 62295274 | -815 | OSBPL2 | 1100 | STRANDS=+-:127 | Validated |
| 20 | 41184282 | 41184388 | -106 | ZHX3 | 449.6 | STRANDS=+-:62 | Validated |
| 1 | 86562531 | 86573603 | -11072 | CLCA4 | 1673 | STRANDS=+-:180 | Validated |
| 16 | 27325117 | 27339881 | -14764 | IL4R | 608.4 | STRANDS=+-:113 | Validated |
| 16 | 83162542 | 83176121 | -13579 | CDH13 | 1358 | STRANDS=+-:252 | Validated |
| X | 85087992 | 85088221 | -229 | APOOL | 747.7 | STRANDS=+-:23 | Validation in only 7/16, dropped |

### Supplemental Table S3. SNPs in deletions overlapping with GTEx expression data.

| gene | SNP | p-value | nes | tissue |
|---|---|---|---|---|
| CLCA4 | rs10489695 | 4.50E-08 | -0.34 | Brain_Caudate(basalganglia) |
| CLCA4 | rs10489695 | 2.2E-05 | -0.27 | Brain_Putamen(basalganglia) |
| CLCA4 | rs11158402 | 2.40E-07 | -0.61 | Brain_Cortex |
| CLCA4 | rs11158402 | 5.50E-07 | -0.8 | Brain_Caudate(basalganglia) |
| CLCA4 | rs11158402 | 4E-06 | -0.66 | Brain_Putamen(basalganglia) |
| CLCA4 | rs11158402 | 6E-06 | -0.92 | Brain_Anteriorcingulatecortex( |
| CLCA4 | rs11158402 | 1.6E-05 | -0.74 | Brain_Amygdala |
| CLCA4 | rs11158402 | 1.7E-05 | -0.7 | Brain_Hypothalamus |
| CLCA4 | rs11404430 | 3.90E-08 | -0.81 | Brain_Putamen(basalganglia) |
| CLCA4 | rs11404430 | 4.30E-07 | -0.85 | Brain_Caudate(basalganglia) |
| CLCA4 | rs11404430 | 8.60E-07 | -0.63 | Brain_Cortex |
| CLCA4 | rs11404430 | 2.2E-06 | -0.85 | Brain_Amygdala |
| CLCA4 | rs11404430 | 6E-06 | -0.92 | Brain_Anteriorcingulatecortex( |
| CLCA4 | rs11404430 | 1.7E-05 | -0.7 | Brain_Hypothalamus |
| CLCA4 | rs14609959 | 3.90E-08 | -0.81 | Brain_Putamen(basalganglia) |
| CLCA4 | rs14609959 | 4.30E-07 | -0.85 | Brain_Caudate(basalganglia) |
| CLCA4 | rs14609959 | 8.60E-07 | -0.63 | Brain_Cortex |
| CLCA4 | rs14609959 | 2.2E-06 | -0.85 | Brain_Amygdala |
| CLCA4 | rs14609959 | 6E-06 | -0.92 | Brain_Anteriorcingulatecortex( |
| CLCA4 | rs14609959 | 1.7E-05 | -0.7 | Brain_Hypothalamus |
| CLCA4 | rs15010112 | 3.90E-08 | -0.81 | Brain_Putamen(basalganglia) |
| CLCA4 | rs15010112 | 4.30E-07 | -0.85 | Brain_Caudate(basalganglia) |
| CLCA4 | rs15010112 | 8.60E-07 | -0.63 | Brain_Cortex |
| CLCA4 | rs15010112 | 2.2E-06 | -0.85 | Brain_Amygdala |
| CLCA4 | rs15010112 | 6E-06 | -0.92 | Brain_Anteriorcingulatecortex( |
| CLCA4 | rs15010112 | 1.7E-05 | -0.7 | Brain_Hypothalamus |
| CLCA4 | rs2231585 | 3.90E-08 | -0.81 | Brain_Putamen(basalganglia) |
| CLCA4 | rs2231585 | 4.30E-07 | -0.85 | Brain_Caudate(basalganglia) |
| CLCA4 | rs2231585 | 8.60E-07 | -0.63 | Brain_Cortex |
| CLCA4 | rs2231585 | 2.2E-06 | -0.85 | Brain_Amygdala |
| CLCA4 | rs2231585 | 6E-06 | -0.92 | Brain_Anteriorcingulatecortex( |
| CLCA4 | rs2231585 | 1.7E-05 | -0.7 | Brain_Hypothalamus |
| CLCA4 | rs2231596 | 2.40E-07 | -0.61 | Brain_Cortex |
| CLCA4 | rs2231596 | 5.50E-07 | -0.8 | Brain_Caudate(basalganglia) |
| CLCA4 | rs2231596 | 4E-06 | -0.66 | Brain_Putamen(basalganglia) |
| CLCA4 | rs2231596 | 6E-06 | -0.92 | Brain_Anteriorcingulatecortex( |
| CLCA4 | rs2231596 | 1.6E-05 | -0.74 | Brain_Amygdala |
| CLCA4 | rs2231596 | 1.7E-05 | -0.7 | Brain_Hypothalamus |
| CLCA4 | rs55660984 | 3.90E-08 | -0.81 | Brain_Putamen(basalganglia) |
| CLCA4 | rs55660984 | 4.30E-07 | -0.85 | Brain_Caudate(basalganglia) |
| CLCA4 | rs55660984 | 8.60E-07 | -0.63 | Brain_Cortex |
| CLCA4 | rs55660984 | 2.2E-06 | -0.85 | Brain_Amygdala |
| CLCA4 | rs55660984 | 6E-06 | -0.92 | Brain_Anteriorcingulatecortex( |
| CLCA4 | rs55660984 | 1.7E-05 | -0.7 | Brain_Hypothalamus |

| | | | | |
|---|---|---|---|---|
| CLCA4 | rs5775914 | 0.00004 | 0.23 | Brain_Caudate(basalganglia) |
| CLCA4 | rs772603 | 1.2E-05 | 0.22 | Brain_Caudate(basalganglia) |
| CLCA4 | rs80245293 | 3.90E-08 | -0.81 | Brain_Putamen(basalganglia) |
| CLCA4 | rs80245293 | 4.30E-07 | -0.85 | Brain_Caudate(basalganglia) |
| CLCA4 | rs80245293 | 8.60E-07 | -0.63 | Brain_Cortex |
| CLCA4 | rs80245293 | 2.2E-06 | -0.85 | Brain_Amygdala |
| CLCA4 | rs80245293 | 6E-06 | -0.92 | Brain_Anteriorcingulatecortex( |
| CLCA4 | rs80245293 | 1.7E-05 | -0.7 | Brain_Hypothalamus |
| FBXO36 | rs11420388 | 2.10E-07 | 0.21 | Adipose_Subcutaneous |
| FBXO36 | rs11420388 | 1.1E-05 | 0.22 | Adipose_Visceral(Omentum) |
| FBXO36 | rs11420388 | 1.20E-10 | 0.38 | Artery_Aorta |
| FBXO36 | rs11420388 | 4.6E-06 | 0.4 | Artery_Coronary |
| FBXO36 | rs11420388 | 4.40E-08 | 0.26 | Artery_Tibial |
| FBXO36 | rs11420388 | 1.6E-06 | 0.27 | Cells_Culturedfibroblasts |
| FBXO36 | rs11420388 | 4.9E-06 | 0.27 | Colon_Transverse |
| FBXO36 | rs11420388 | 8.20E-08 | 0.35 | Esophagus_GastroesophagealJunc |
| FBXO36 | rs11420388 | 7.30E-07 | 0.27 | Esophagus_Muscularis |
| FBXO36 | rs11420388 | 3.1E-06 | 0.31 | Heart_AtrialAppendage |
| FBXO36 | rs11420388 | 4.3E-06 | 0.22 | Nerve_Tibial |
| FBXO36 | rs11420388 | 3.4E-06 | 0.21 | Skin_NotSunExposed(Suprapubic) |
| FBXO36 | rs11420388 | 0.00006 | 0.16 | Skin_SunExposed(Lowerleg) |
| FBXO36 | rs11420388 | 2.3E-06 | 0.18 | Thyroid |
| FBXO36 | rs11683062 | 2.3E-06 | -0.21 | Artery_Aorta |
| FBXO36 | rs11683062 | 1.20E-07 | -0.19 | Artery_Tibial |
| FBXO36 | rs11683062 | 8.60E-09 | -0.23 | Cells_Culturedfibroblasts |
| FBXO36 | rs11683062 | 4.4E-05 | -0.14 | Esophagus_Mucosa |
| FBXO36 | rs1660718 | 4.9E-05 | -0.19 | Cells_Culturedfibroblasts |
| FBXO36 | rs2162524 | 5.10E-09 | 0.23 | Cells_Culturedfibroblasts |
| FBXO36 | rs2433733 | 3.40E-09 | -0.23 | Cells_Culturedfibroblasts |
| FBXO36 | rs35547214 | 2.60E-09 | 0.24 | Cells_Culturedfibroblasts |
| FBXO36 | rs3856526 | 1.50E-07 | 0.21 | Adipose_Subcutaneous |
| FBXO36 | rs3856526 | 5E-06 | 0.23 | Adipose_Visceral(Omentum) |
| FBXO36 | rs3856526 | 1.20E-10 | 0.38 | Artery_Aorta |
| FBXO36 | rs3856526 | 4.6E-06 | 0.4 | Artery_Coronary |
| FBXO36 | rs3856526 | 3.60E-08 | 0.26 | Artery_Tibial |
| FBXO36 | rs3856526 | 1.5E-06 | 0.27 | Cells_Culturedfibroblasts |
| FBXO36 | rs3856526 | 4.9E-06 | 0.27 | Colon_Transverse |
| FBXO36 | rs3856526 | 7.10E-08 | 0.36 | Esophagus_GastroesophagealJunc |
| FBXO36 | rs3856526 | 7.30E-07 | 0.27 | Esophagus_Muscularis |
| FBXO36 | rs3856526 | 3.1E-06 | 0.31 | Heart_AtrialAppendage |
| FBXO36 | rs3856526 | 4.3E-06 | 0.22 | Nerve_Tibial |
| FBXO36 | rs3856526 | 3.5E-06 | 0.21 | Skin_NotSunExposed(Suprapubic) |
| FBXO36 | rs3856526 | 0.00006 | 0.16 | Skin_SunExposed(Lowerleg) |
| FBXO36 | rs3856526 | 2.3E-06 | 0.18 | Thyroid |
| FBXO36 | rs536707 | 2.9E-06 | -0.13 | Adipose_Subcutaneous |
| FBXO36 | rs536707 | 6.30E-08 | -0.19 | Adipose_Visceral(Omentum) |
| FBXO36 | rs536707 | 1.50E-11 | -0.28 | Artery_Aorta |

| | | | | |
|---|---|---|---|---|
| FBXO36 | rs536707 | 2.30E-11 | -0.22 | Artery_Tibial |
| FBXO36 | rs536707 | 4.3E-06 | -0.16 | Breast_MammaryTissue |
| FBXO36 | rs536707 | 1.10E-20 | -0.34 | Cells_Culturedfibroblasts |
| FBXO36 | rs536707 | 8.60E-09 | -0.27 | Colon_Sigmoid |
| FBXO36 | rs536707 | 1.5E-05 | -0.17 | Colon_Transverse |
| FBXO36 | rs536707 | 2.20E-07 | -0.25 | Esophagus_GastroesophagealJunc |
| FBXO36 | rs536707 | 7.20E-07 | -0.2 | Esophagus_Muscularis |
| FBXO36 | rs536707 | 4.00E-09 | -0.27 | Heart_AtrialAppendage |
| FBXO36 | rs536707 | 9.00E-07 | -0.27 | Heart_LeftVentricle |
| FBXO36 | rs536707 | 0.00001 | -0.26 | Ovary |
| FBXO36 | rs536707 | 1.00E-08 | -0.29 | Pituitary |
| FBXO36 | rs536707 | 1.5E-05 | -0.11 | Thyroid |
| FBXO36 | rs553536 | 8.10E-07 | -0.19 | Adipose_Subcutaneous |
| FBXO36 | rs553536 | 5.6E-06 | -0.22 | Adipose_Visceral(Omentum) |
| FBXO36 | rs553536 | 4.80E-10 | -0.36 | Artery_Aorta |
| FBXO36 | rs553536 | 4.2E-06 | -0.39 | Artery_Coronary |
| FBXO36 | rs553536 | 3.00E-08 | -0.25 | Artery_Tibial |
| FBXO36 | rs553536 | 2.4E-05 | -0.29 | Brain_Hypothalamus |
| FBXO36 | rs553536 | 5.9E-06 | -0.25 | Cells_Culturedfibroblasts |
| FBXO36 | rs553536 | 2.8E-06 | -0.26 | Colon_Transverse |
| FBXO36 | rs553536 | 2.10E-07 | -0.34 | Esophagus_GastroesophagealJunc |
| FBXO36 | rs553536 | 2.8E-06 | -0.25 | Esophagus_Muscularis |
| FBXO36 | rs553536 | 1.1E-05 | -0.21 | Nerve_Tibial |
| FBXO36 | rs553536 | 1.1E-06 | -0.22 | Skin_NotSunExposed(Suprapubic) |
| FBXO36 | rs553536 | 5.8E-06 | -0.17 | Thyroid |
| FBXO36 | rs55953045 | 6.00E-17 | -0.72 | Adipose_Subcutaneous |
| FBXO36 | rs55953045 | 2.20E-11 | -0.74 | Adipose_Visceral(Omentum) |
| FBXO36 | rs55953045 | 2.10E-08 | -0.91 | AdrenalGland |
| FBXO36 | rs55953045 | 7.50E-10 | -0.78 | Artery_Aorta |
| FBXO36 | rs55953045 | 3.40E-17 | -0.94 | Artery_Tibial |
| FBXO36 | rs55953045 | 2.70E-11 | -1.2 | Brain_Amygdala |
| FBXO36 | rs55953045 | 6.10E-11 | -1.1 | Brain_Anteriorcingulatecortex( |
| FBXO36 | rs55953045 | 6.50E-14 | -1.3 | Brain_Caudate(basalganglia) |
| FBXO36 | rs55953045 | 9.10E-19 | -1.6 | Brain_CerebellarHemisphere |
| FBXO36 | rs55953045 | 1.70E-07 | -1.1 | Brain_Cerebellum |
| FBXO36 | rs55953045 | 6.50E-19 | -1.9 | Brain_Cortex |
| FBXO36 | rs55953045 | 6.00E-12 | -1.3 | Brain_FrontalCortex(BA9) |
| FBXO36 | rs55953045 | 1.40E-11 | -1.2 | Brain_Hippocampus |
| FBXO36 | rs55953045 | 1.40E-12 | -0.99 | Brain_Hypothalamus |
| FBXO36 | rs55953045 | 1.50E-16 | -1.4 | Brain_Nucleusaccumbens(basalga |
| FBXO36 | rs55953045 | 8.80E-16 | -1.6 | Brain_Putamen(basalganglia) |
| FBXO36 | rs55953045 | 5.50E-09 | -1.3 | Brain_Spinalcord(cervicalc-1) |
| FBXO36 | rs55953045 | 9.30E-11 | -1.5 | Brain_Substantianigra |
| FBXO36 | rs55953045 | 1.10E-10 | -0.7 | Breast_MammaryTissue |
| FBXO36 | rs55953045 | 4.3E-06 | -0.59 | Cells_Culturedfibroblasts |
| FBXO36 | rs55953045 | 2.3E-05 | -0.58 | Colon_Sigmoid |
| FBXO36 | rs55953045 | 4.80E-09 | -0.79 | Colon_Transverse |

| | | | | |
|---|---|---|---|---|
| FBXO36 | rs55953045 | 1.50E-07 | -0.9 | Esophagus_GastroesophagealJunc |
| FBXO36 | rs55953045 | 1.90E-08 | -0.61 | Esophagus_Mucosa |
| FBXO36 | rs55953045 | 9.90E-14 | -0.88 | Esophagus_Muscularis |
| FBXO36 | rs55953045 | 4.70E-07 | -0.74 | Heart_AtrialAppendage |
| FBXO36 | rs55953045 | 8.80E-12 | -0.6 | Lung |
| FBXO36 | rs55953045 | 1.10E-20 | -0.97 | Nerve_Tibial |
| FBXO36 | rs55953045 | 5.80E-07 | -0.8 | Pituitary |
| FBXO36 | rs55953045 | 7.50E-17 | -0.83 | Skin_NotSunExposed(Suprapubic) |
| FBXO36 | rs55953045 | 3.40E-17 | -0.78 | Skin_SunExposed(Lowerleg) |
| FBXO36 | rs55953045 | 4.10E-07 | -0.81 | Stomach |
| FBXO36 | rs55953045 | 1.50E-13 | -0.63 | Thyroid |
| FBXO36 | rs62191699 | 1.50E-09 | 0.24 | Cells_Culturedfibroblasts |
| FBXO36 | rs62191702 | 1.50E-09 | 0.24 | Cells_Culturedfibroblasts |
| FBXO36 | rs6722477 | 4.80E-07 | 0.2 | Adipose_Subcutaneous |
| FBXO36 | rs6722477 | 2.3E-05 | 0.21 | Adipose_Visceral(Omentum) |
| FBXO36 | rs6722477 | 2.70E-10 | 0.37 | Artery_Aorta |
| FBXO36 | rs6722477 | 4.6E-06 | 0.4 | Artery_Coronary |
| FBXO36 | rs6722477 | 8.90E-08 | 0.25 | Artery_Tibial |
| FBXO36 | rs6722477 | 2.3E-06 | 0.26 | Cells_Culturedfibroblasts |
| FBXO36 | rs6722477 | 7.6E-06 | 0.26 | Colon_Transverse |
| FBXO36 | rs6722477 | 8.20E-08 | 0.35 | Esophagus_GastroesophagealJunc |
| FBXO36 | rs6722477 | 1.5E-06 | 0.26 | Esophagus_Muscularis |
| FBXO36 | rs6722477 | 1.7E-05 | 0.29 | Heart_AtrialAppendage |
| FBXO36 | rs6722477 | 1.5E-05 | 0.21 | Nerve_Tibial |
| FBXO36 | rs6722477 | 4E-06 | 0.21 | Skin_NotSunExposed(Suprapubic) |
| FBXO36 | rs6722477 | 0.00012 | 0.15 | Skin_SunExposed(Lowerleg) |
| FBXO36 | rs6722477 | 5.8E-06 | 0.17 | Thyroid |
| FBXO36 | rs72049645 | 8.2E-06 | -0.18 | Adipose_Subcutaneous |
| FBXO36 | rs72049645 | 7.4E-05 | -0.24 | Artery_Aorta |
| FBXO36 | rs72049645 | 1.8E-06 | -0.23 | Artery_Tibial |
| FBXO36 | rs72049645 | 6.20E-07 | -0.51 | Brain_Cortex |
| FBXO36 | rs72049645 | 4.5E-05 | -0.16 | Thyroid |
| FBXO36 | rs73103553 | 1.50E-07 | 0.21 | Adipose_Subcutaneous |
| FBXO36 | rs73103553 | 5E-06 | 0.23 | Adipose_Visceral(Omentum) |
| FBXO36 | rs73103553 | 1.20E-10 | 0.38 | Artery_Aorta |
| FBXO36 | rs73103553 | 4.6E-06 | 0.4 | Artery_Coronary |
| FBXO36 | rs73103553 | 3.60E-08 | 0.26 | Artery_Tibial |
| FBXO36 | rs73103553 | 1.5E-06 | 0.27 | Cells_Culturedfibroblasts |
| FBXO36 | rs73103553 | 4.9E-06 | 0.27 | Colon_Transverse |
| FBXO36 | rs73103553 | 7.10E-08 | 0.36 | Esophagus_GastroesophagealJunc |
| FBXO36 | rs73103553 | 7.30E-07 | 0.27 | Esophagus_Muscularis |
| FBXO36 | rs73103553 | 3.1E-06 | 0.31 | Heart_AtrialAppendage |
| FBXO36 | rs73103553 | 4.3E-06 | 0.22 | Nerve_Tibial |
| FBXO36 | rs73103553 | 3.5E-06 | 0.21 | Skin_NotSunExposed(Suprapubic) |
| FBXO36 | rs73103553 | 0.00006 | 0.16 | Skin_SunExposed(Lowerleg) |
| FBXO36 | rs73103553 | 2.3E-06 | 0.18 | Thyroid |
| FBXO36 | rs75277969 | 6.00E-17 | -0.72 | Adipose_Subcutaneous |

| | | | | |
|---|---|---|---|---|
| FBXO36 | rs75277969 | 2.20E-11 | -0.74 | Adipose_Visceral(Omentum) |
| FBXO36 | rs75277969 | 2.10E-08 | -0.91 | AdrenalGland |
| FBXO36 | rs75277969 | 7.50E-10 | -0.78 | Artery_Aorta |
| FBXO36 | rs75277969 | 3.40E-17 | -0.94 | Artery_Tibial |
| FBXO36 | rs75277969 | 2.70E-11 | -1.2 | Brain_Amygdala |
| FBXO36 | rs75277969 | 6.10E-11 | -1.1 | Brain_Anteriorcingulatecortex( |
| FBXO36 | rs75277969 | 6.50E-14 | -1.3 | Brain_Caudate(basalganglia) |
| FBXO36 | rs75277969 | 9.10E-19 | -1.6 | Brain_CerebellarHemisphere |
| FBXO36 | rs75277969 | 1.70E-07 | -1.1 | Brain_Cerebellum |
| FBXO36 | rs75277969 | 6.50E-19 | -1.9 | Brain_Cortex |
| FBXO36 | rs75277969 | 6.00E-12 | -1.3 | Brain_FrontalCortex(BA9) |
| FBXO36 | rs75277969 | 1.40E-11 | -1.2 | Brain_Hippocampus |
| FBXO36 | rs75277969 | 1.40E-12 | -0.99 | Brain_Hypothalamus |
| FBXO36 | rs75277969 | 1.50E-16 | -1.4 | Brain_Nucleusaccumbens(basalga |
| FBXO36 | rs75277969 | 8.80E-16 | -1.6 | Brain_Putamen(basalganglia) |
| FBXO36 | rs75277969 | 5.50E-09 | -1.3 | Brain_Spinalcord(cervicalc-1) |
| FBXO36 | rs75277969 | 9.30E-11 | -1.5 | Brain_Substantianigra |
| FBXO36 | rs75277969 | 1.10E-10 | -0.7 | Breast_MammaryTissue |
| FBXO36 | rs75277969 | 4.3E-06 | -0.59 | Cells_Culturedfibroblasts |
| FBXO36 | rs75277969 | 2.3E-05 | -0.58 | Colon_Sigmoid |
| FBXO36 | rs75277969 | 4.80E-09 | -0.79 | Colon_Transverse |
| FBXO36 | rs75277969 | 1.50E-07 | -0.9 | Esophagus_GastroesophagealJunc |
| FBXO36 | rs75277969 | 1.90E-08 | -0.61 | Esophagus_Mucosa |
| FBXO36 | rs75277969 | 9.90E-14 | -0.88 | Esophagus_Muscularis |
| FBXO36 | rs75277969 | 4.70E-07 | -0.74 | Heart_AtrialAppendage |
| FBXO36 | rs75277969 | 8.80E-12 | -0.6 | Lung |
| FBXO36 | rs75277969 | 1.10E-20 | -0.97 | Nerve_Tibial |
| FBXO36 | rs75277969 | 5.80E-07 | -0.8 | Pituitary |
| FBXO36 | rs75277969 | 7.50E-17 | -0.83 | Skin_NotSunExposed(Suprapubic) |
| FBXO36 | rs75277969 | 3.40E-17 | -0.78 | Skin_SunExposed(Lowerleg) |
| FBXO36 | rs75277969 | 4.10E-07 | -0.81 | Stomach |
| FBXO36 | rs75277969 | 1.50E-13 | -0.63 | Thyroid |
| FBXO36 | rs7558844 | 1.9E-05 | 0.66 | Testis |
| FBXO36 | rs7605745 | 2.10E-07 | 0.21 | Adipose_Subcutaneous |
| FBXO36 | rs7605745 | 7.9E-06 | 0.22 | Adipose_Visceral(Omentum) |
| FBXO36 | rs7605745 | 1.20E-10 | 0.38 | Artery_Aorta |
| FBXO36 | rs7605745 | 4.6E-06 | 0.4 | Artery_Coronary |
| FBXO36 | rs7605745 | 4.00E-08 | 0.26 | Artery_Tibial |
| FBXO36 | rs7605745 | 1.6E-06 | 0.27 | Cells_Culturedfibroblasts |
| FBXO36 | rs7605745 | 9E-06 | 0.26 | Colon_Transverse |
| FBXO36 | rs7605745 | 7.10E-08 | 0.36 | Esophagus_GastroesophagealJunc |
| FBXO36 | rs7605745 | 9.20E-07 | 0.27 | Esophagus_Muscularis |
| FBXO36 | rs7605745 | 1.7E-05 | 0.29 | Heart_AtrialAppendage |
| FBXO36 | rs7605745 | 1.5E-05 | 0.21 | Nerve_Tibial |
| FBXO36 | rs7605745 | 5.3E-06 | 0.21 | Skin_NotSunExposed(Suprapubic) |
| FBXO36 | rs7605745 | 7.6E-05 | 0.16 | Skin_SunExposed(Lowerleg) |
| FBXO36 | rs7605745 | 3E-06 | 0.18 | Thyroid |

| | | | | |
|---|---|---|---|---|
| FBXO36 | rs77408380 | 7.90E-07 | 0.22 | Adipose_Subcutaneous |
| FBXO36 | rs77408380 | 3.30E-08 | 0.35 | Artery_Aorta |
| FBXO36 | rs77408380 | 1.7E-05 | 0.4 | Artery_Coronary |
| FBXO36 | rs77408380 | 3.80E-07 | 0.26 | Artery_Tibial |
| FBXO36 | rs77408380 | 0.00008 | 0.25 | Cells_Culturedfibroblasts |
| FBXO36 | rs77408380 | 6.1E-06 | 0.29 | Colon_Transverse |
| FBXO36 | rs77408380 | 7.60E-08 | 0.38 | Esophagus_GastroesophagealJunc |
| FBXO36 | rs77408380 | 1.2E-05 | 0.26 | Esophagus_Muscularis |
| FBXO36 | rs77408380 | 2.8E-05 | 0.3 | Heart_AtrialAppendage |
| FBXO36 | rs77408380 | 6.8E-05 | 0.21 | Nerve_Tibial |
| FBXO36 | rs77408380 | 4.6E-05 | 0.2 | Skin_NotSunExposed(Suprapubic) |
| FBXO36 | rs77408380 | 4.7E-05 | 0.17 | Thyroid |
| IL4R | rs12445247 | 0.000011 | 0.19 | AdrenalGland |
| IL4R | rs12445247 | 0.00001 | -0.28 | Brain_Cerebellum |
| IL4R | rs12445247 | 0.000045 | 0.098 | Esophagus_Mucosa |
| IL4R | rs12445247 | 5.90E-07 | 0.1 | Lung |
| IL4R | rs12445247 | 4.20E-14 | 0.12 | WholeBlood |
| IL4R | rs12708699 | 0.0000025 | -0.14 | Adipose_Visceral(Omentum) |
| IL4R | rs12708700 | 0.0000025 | -0.14 | Adipose_Visceral(Omentum) |
| IL4R | rs12925861 | 1.40E-09 | -0.28 | AdrenalGland |
| IL4R | rs12925861 | 0.000015 | 0.31 | Brain_CerebellarHemisphere |
| IL4R | rs12925861 | 0.000007 | 0.3 | Brain_Cerebellum |
| IL4R | rs12925861 | 3.10E-08 | -0.15 | Cells_Culturedfibroblasts |
| IL4R | rs12925861 | 6.60E-13 | -0.12 | WholeBlood |
| IL4R | rs18774430 | 0.000054 | -0.13 | Thyroid |
| IL4R | rs1981551 | 5.80E-12 | -0.32 | AdrenalGland |
| IL4R | rs1981551 | 0.0000022 | 0.34 | Brain_CerebellarHemisphere |
| IL4R | rs1981551 | 0.0000051 | 0.31 | Brain_Cerebellum |
| IL4R | rs1981551 | 7.00E-08 | -0.15 | Cells_Culturedfibroblasts |
| IL4R | rs1981551 | 0.00001 | -0.18 | Liver |
| IL4R | rs1981551 | 0.0000064 | -0.16 | Pancreas |
| IL4R | rs1981551 | 6.10E-14 | -0.13 | WholeBlood |
| IL4R | rs20003778 | 0.000054 | -0.13 | Thyroid |
| IL4R | rs2107354 | 7.90E-10 | -0.28 | AdrenalGland |
| IL4R | rs2107354 | 0.000022 | 0.3 | Brain_CerebellarHemisphere |
| IL4R | rs2107354 | 0.000017 | 0.29 | Brain_Cerebellum |
| IL4R | rs2107354 | 1.10E-07 | -0.14 | Cells_Culturedfibroblasts |
| IL4R | rs2107354 | 3.20E-13 | -0.12 | WholeBlood |
| IL4R | rs2107355 | 1.50E-10 | -0.29 | AdrenalGland |
| IL4R | rs2107355 | 0.000011 | 0.31 | Brain_CerebellarHemisphere |
| IL4R | rs2107355 | 0.0000086 | 0.3 | Brain_Cerebellum |
| IL4R | rs2107355 | 0.0000025 | -0.13 | Cells_Culturedfibroblasts |
| IL4R | rs2107355 | 0.0000065 | -0.18 | Liver |
| IL4R | rs2107355 | 0.000016 | -0.15 | Pancreas |
| IL4R | rs2107355 | 2.30E-10 | -0.11 | WholeBlood |
| IL4R | rs2382720 | 0.000008 | -0.19 | AdrenalGland |
| IL4R | rs2382720 | 0.0000049 | 0.28 | Brain_Cerebellum |

| | | | | |
|---|---|---|---|---|
| IL4R | rs2382720 | 0.0000054 | -0.091 | Lung |
| IL4R | rs2382720 | 5.40E-08 | -0.082 | WholeBlood |
| IL4R | rs3024530 | 0.000025 | -0.18 | AdrenalGland |
| IL4R | rs3024530 | 0.000099 | -0.092 | Esophagus_Mucosa |
| IL4R | rs3024530 | 2.80E-07 | -0.099 | Lung |
| IL4R | rs3024530 | 1.90E-07 | -0.078 | WholeBlood |
| IL4R | rs34724561 | 0.000028 | -0.18 | AdrenalGland |
| IL4R | rs34724561 | 0.0000096 | 0.27 | Brain_Cerebellum |
| IL4R | rs34724561 | 0.0000029 | -0.094 | Lung |
| IL4R | rs34724561 | 2.40E-07 | -0.078 | WholeBlood |
| IL4R | rs34943813 | 0.000054 | -0.13 | Thyroid |
| IL4R | rs35004258 | 1.20E-11 | -0.31 | AdrenalGland |
| IL4R | rs35004258 | 0.000019 | 0.29 | Brain_Cerebellum |
| IL4R | rs35004258 | 1.00E-08 | -0.16 | Cells_Culturedfibroblasts |
| IL4R | rs35004258 | 0.0000063 | -0.15 | Pancreas |
| IL4R | rs35004258 | 2.10E-16 | -0.14 | WholeBlood |
| IL4R | rs3785356 | 1.20E-10 | -0.3 | AdrenalGland |
| IL4R | rs3785356 | 0.0000095 | 0.33 | Brain_CerebellarHemisphere |
| IL4R | rs3785356 | 0.0000056 | 0.32 | Brain_Cerebellum |
| IL4R | rs3785356 | 1.60E-07 | -0.15 | Cells_Culturedfibroblasts |
| IL4R | rs3785356 | 0.00001 | -0.19 | Liver |
| IL4R | rs3785356 | 0.0000047 | -0.16 | Pancreas |
| IL4R | rs3785356 | 7.00E-13 | -0.12 | WholeBlood |
| IL4R | rs3916997 | 0.0000019 | -0.21 | AdrenalGland |
| IL4R | rs3916997 | 0.0000027 | 0.28 | Brain_Cerebellum |
| IL4R | rs3916997 | 0.0001 | -0.092 | Esophagus_Mucosa |
| IL4R | rs3916997 | 0.0000078 | -0.089 | Lung |
| IL4R | rs3916997 | 7.10E-08 | -0.082 | WholeBlood |
| IL4R | rs4787948 | 6.30E-10 | -0.28 | AdrenalGland |
| IL4R | rs4787948 | 0.000043 | 0.3 | Brain_CerebellarHemisphere |
| IL4R | rs4787948 | 1.90E-08 | -0.15 | Cells_Culturedfibroblasts |
| IL4R | rs4787948 | 0.000032 | -0.14 | Pancreas |
| IL4R | rs4787948 | 1.20E-15 | -0.13 | WholeBlood |
| IL4R | rs4787951 | 9.70E-12 | -0.31 | AdrenalGland |
| IL4R | rs4787951 | 7.60E-07 | 0.35 | Brain_CerebellarHemisphere |
| IL4R | rs4787951 | 0.0000034 | 0.31 | Brain_Cerebellum |
| IL4R | rs4787951 | 1.60E-07 | -0.15 | Cells_Culturedfibroblasts |
| IL4R | rs4787951 | 0.000011 | -0.18 | Liver |
| IL4R | rs4787951 | 0.000018 | -0.15 | Pancreas |
| IL4R | rs4787951 | 9.90E-12 | -0.12 | WholeBlood |
| IL4R | rs56943160 | 0.000028 | 0.29 | Heart_AtrialAppendage |
| IL4R | rs56943160 | 0.0000015 | 0.34 | Heart_LeftVentricle |
| IL4R | rs58653621 | 0.0000013 | -0.21 | AdrenalGland |
| IL4R | rs58653621 | 0.0000079 | 0.27 | Brain_Cerebellum |
| IL4R | rs58653621 | 0.000091 | -0.094 | Esophagus_Mucosa |
| IL4R | rs58653621 | 0.000016 | -0.086 | Lung |
| IL4R | rs58653621 | 9.20E-09 | -0.087 | WholeBlood |

| | | | | |
|---|---|---|---|---|
| IL4R | rs61373663 | 0.000028 | 0.29 | Heart_AtrialAppendage |
| IL4R | rs61373663 | 0.0000015 | 0.34 | Heart_LeftVentricle |
| IL4R | rs6498013 | 7.10E-10 | -0.28 | AdrenalGland |
| IL4R | rs6498013 | 0.000023 | 0.28 | Brain_Cerebellum |
| IL4R | rs6498013 | 1.30E-08 | -0.16 | Cells_Culturedfibroblasts |
| IL4R | rs6498013 | 5.10E-16 | -0.14 | WholeBlood |
| IL4R | rs71388093 | 3.70E-12 | -0.32 | AdrenalGland |
| IL4R | rs71388093 | 0.000053 | 0.29 | Brain_CerebellarHemisphere |
| IL4R | rs71388093 | 0.000026 | 0.28 | Brain_Cerebellum |
| IL4R | rs71388093 | 2.80E-08 | -0.15 | Cells_Culturedfibroblasts |
| IL4R | rs71388093 | 0.000019 | -0.18 | Liver |
| IL4R | rs71388093 | 0.0000062 | -0.16 | Pancreas |
| IL4R | rs71388093 | 1.60E-17 | -0.14 | WholeBlood |
| IL4R | rs7190472 | 4.80E-10 | -0.29 | AdrenalGland |
| IL4R | rs7190472 | 0.000023 | 0.31 | Brain_CerebellarHemisphere |
| IL4R | rs7190472 | 0.000014 | 0.29 | Brain_Cerebellum |
| IL4R | rs7190472 | 2.70E-08 | -0.15 | Cells_Culturedfibroblasts |
| IL4R | rs7190472 | 2.00E-13 | -0.12 | WholeBlood |
| IL4R | rs9673499 | 9.00E-12 | -0.31 | AdrenalGland |
| IL4R | rs9673499 | 1.80E-08 | -0.16 | Cells_Culturedfibroblasts |
| IL4R | rs9673499 | 0.0000092 | -0.19 | Liver |
| IL4R | rs9673499 | 0.0000015 | -0.16 | Pancreas |
| IL4R | rs9673499 | 1.10E-17 | -0.14 | WholeBlood |
| IL4R | rs9929928 | 0.000034 | -0.12 | Adipose_Visceral(Omentum) |
| IL4R | rs9940480 | 0.0000029 | -0.2 | AdrenalGland |
| IL4R | rs9940480 | 0.000012 | 0.27 | Brain_Cerebellum |
| IL4R | rs9940480 | 0.0000023 | -0.094 | Lung |
| IL4R | rs9940480 | 1.30E-07 | -0.079 | WholeBlood |
| LMNTD1 | rs11048077 | 7.00E-07 | -0.14 | Testis |
| LMNTD1 | rs1479509 | 9.80E-07 | -0.13 | Testis |
| LMNTD1 | rs2169990 | 1.40E-07 | -0.17 | Testis |
| LMNTD1 | rs7968985 | 7.00E-07 | -0.14 | Testis |
| MPST | rs10427747 | 5.70E-08 | 0.18 | Muscle_Skeletal |
| MPST | rs10427747 | 1.20E-16 | 0.51 | Testis |
| MPST | rs10427757 | 1.50E-07 | 0.32 | Testis |
| MPST | rs10427778 | 0.000053 | 0.14 | Muscle_Skeletal |
| MPST | rs10427778 | 1.20E-07 | 0.33 | Testis |
| MPST | rs11704682 | 3.20E-07 | 0.47 | Brain_Spinalcord(cervicalc-1) |
| MPST | rs11704682 | 8.20E-24 | -0.28 | Colon_Transverse |
| MPST | rs11704682 | 1.80E-09 | -0.38 | Liver |
| MPST | rs11704682 | 5.10E-09 | -0.24 | SmallIntestine_TerminalIleum |
| MPST | rs12158886 | 0.000026 | -0.61 | Brain_Cerebellum |
| MPST | rs12158886 | 2.70E-07 | -0.8 | Brain_Cortex |
| MPST | rs12158886 | 0.000028 | -0.52 | Brain_Nucleusaccumbens(basalga |
| MPST | rs12158886 | 0.000098 | -0.32 | Muscle_Skeletal |
| MPST | rs12158886 | 4.80E-08 | -0.74 | Testis |
| MPST | rs12158886 | 3.40E-08 | 0.56 | WholeBlood |

| | | | | |
|---|---|---|---|---|
| MPST | rs4821585 | 5.20E-07 | 0.2 | Brain_Caudate(basalganglia) |
| MPST | rs4821585 | 0.0000039 | 0.22 | Brain_Putamen(basalganglia) |
| MPST | rs4821585 | 0.000036 | -0.091 | Colon_Transverse |
| MPST | rs4821585 | 0.0000014 | -0.19 | Liver |
| MPST | rs4821585 | 2.90E-18 | -0.22 | Thyroid |
| MPST | rs57522265 | 0.000003 | -0.3 | Artery_Tibial |
| MPST | rs57522265 | 0.000029 | -0.83 | Brain_Cerebellum |
| MPST | rs57522265 | 5.30E-19 | 0.7 | Cells_Culturedfibroblasts |
| MPST | rs57522265 | 1.60E-07 | 0.38 | Skin_NotSunExposed(Suprapubic) |
| MPST | rs57522265 | 2.40E-14 | 0.62 | WholeBlood |
| MPST | rs5756487 | 4.90E-11 | 0.25 | Brain_Caudate(basalganglia) |
| MPST | rs5756487 | 2.20E-08 | 0.26 | Brain_Putamen(basalganglia) |
| MPST | rs5756487 | 0.0000027 | -0.099 | Colon_Transverse |
| MPST | rs5756487 | 0.0000045 | -0.18 | Liver |
| MPST | rs5756487 | 0.000026 | 0.075 | Muscle_Skeletal |
| MPST | rs5756487 | 2.20E-18 | -0.21 | Thyroid |
| MPST | rs5756488 | 0.000012 | 0.19 | Brain_Caudate(basalganglia) |
| MPST | rs5756488 | 0.0000039 | 0.2 | Ovary |
| MPST | rs5756488 | 6.50E-13 | -0.19 | Thyroid |
| MPST | rs5756489 | 1.30E-10 | 0.25 | Brain_Caudate(basalganglia) |
| MPST | rs5756489 | 8.80E-09 | 0.26 | Brain_Putamen(basalganglia) |
| MPST | rs5756489 | 0.0000028 | -0.099 | Colon_Transverse |
| MPST | rs5756489 | 0.0000055 | -0.18 | Liver |
| MPST | rs5756489 | 0.000027 | 0.076 | Muscle_Skeletal |
| MPST | rs5756489 | 5.40E-20 | -0.22 | Thyroid |
| MPST | rs58029343 | 0.000029 | -0.23 | Artery_Tibial |
| MPST | rs58029343 | 0.000025 | -0.62 | Brain_Caudate(basalganglia) |
| MPST | rs58029343 | 0.0000013 | -0.68 | Brain_Cerebellum |
| MPST | rs58029343 | 0.0000011 | -0.75 | Brain_Cortex |
| MPST | rs58029343 | 0.000012 | -0.51 | Brain_Nucleusaccumbens(basalga |
| MPST | rs58029343 | 9.80E-15 | 0.59 | Cells_Culturedfibroblasts |
| MPST | rs58029343 | 0.0000024 | -0.33 | Muscle_Skeletal |
| MPST | rs58029343 | 3.90E-08 | 0.37 | Skin_NotSunExposed(Suprapubic) |
| MPST | rs58029343 | 6.00E-09 | -0.74 | Testis |
| MPST | rs58029343 | 6.80E-17 | 0.63 | WholeBlood |
| MPST | rs59695783 | 0.000029 | -0.23 | Artery_Tibial |
| MPST | rs59695783 | 0.000025 | -0.62 | Brain_Caudate(basalganglia) |
| MPST | rs59695783 | 0.0000013 | -0.68 | Brain_Cerebellum |
| MPST | rs59695783 | 0.0000011 | -0.75 | Brain_Cortex |
| MPST | rs59695783 | 0.000012 | -0.51 | Brain_Nucleusaccumbens(basalga |
| MPST | rs59695783 | 9.80E-15 | 0.59 | Cells_Culturedfibroblasts |
| MPST | rs59695783 | 0.0000024 | -0.33 | Muscle_Skeletal |
| MPST | rs59695783 | 3.90E-08 | 0.37 | Skin_NotSunExposed(Suprapubic) |
| MPST | rs59695783 | 6.00E-09 | -0.74 | Testis |
| MPST | rs59695783 | 6.80E-17 | 0.63 | WholeBlood |
| MPST | rs62230792 | 3.20E-07 | 0.47 | Brain_Spinalcord(cervicalc-1) |
| MPST | rs62230792 | 8.20E-24 | -0.28 | Colon_Transverse |

| | | | | |
|---|---|---|---|---|
| MPST | rs62230792 | 1.80E-09 | -0.38 | Liver |
| MPST | rs62230792 | 5.10E-09 | -0.24 | SmallIntestine_TerminalIleum |
| OSBPL2 | rs6121980 | 6.50E-07 | 0.45 | Testis |
| PIPOX | rs15071441 | 1.2E-06 | 0.7 | AdrenalGland |
| PIPOX | rs72815754 | 1.2E-06 | 0.7 | AdrenalGland |
| PIPOX | rs72815755 | 1.2E-06 | 0.7 | AdrenalGland |
| PIPOX | rs9912656 | 2.9E-05 | 0.49 | AdrenalGland |
| ZNF44 | rs438963 | 1.70E-12 | 0.4 | Brain_CerebellarHemisphere |
| ZNF44 | rs438963 | 1.6E-06 | 0.31 | Brain_Caudate(basalganglia) |
| ZNF44 | rs438963 | 4E-06 | 0.28 | Brain_Cerebellum |
| ZNF44 | rs438963 | 2.3E-05 | 0.28 | Pituitary |
| ZNF44 | rs438963 | 4.7E-05 | 0.13 | Thyroid |
| ZNF44 | rs533121 | 5.30E-10 | 0.37 | Brain_CerebellarHemisphere |
| ZNF44 | rs533121 | 4.7E-06 | -0.16 | Cells_Culturedfibroblasts |
| ZNF44 | rs533121 | 1.3E-05 | 0.29 | Brain_Caudate(basalganglia) |
| ZNF44 | rs533121 | 8.8E-05 | 0.24 | Brain_Cerebellum |
| ZNF44 | rs533121 | 0.00031 | -0.12 | Skin_SunExposed(Lowerleg) |
| ZNF44 | rs56693009 | 3.6E-05 | -0.18 | WholeBlood |
| ZNF44 | rs56693009 | 0.00008 | 0.32 | Brain_CerebellarHemisphere |
| ZNF44 | rs59305069 | 3.6E-05 | -0.18 | WholeBlood |
| ZNF44 | rs59305069 | 0.00008 | 0.32 | Brain_CerebellarHemisphere |
| ZNF44 | rs59549715 | 0.00014 | -0.18 | WholeBlood |

[1] NES = Normalized effect size: effect of the alternative allele (ALT) relative to the reference allele (REF)
