## Supplemental Informaton for "Intragenic deletions from whole genome sequencing of 1054 suicide deaths"

**Supplemental Information.**

**Image files for In silico representations of deletions**

**File1.** "Samplots.zip" Samplots from SAMPLOT software (png files) for the 11 validated deletions.

**File2.** "Validation images.zip" Visualizations of manual validations from agarose gels and taqman assays for the 11 validated deletions.

File2 Image descriptions:
"chr1_86562530_86573603_CLCA4_Validation.pdf". Taqman visualization from CopyCaller. Bars 1-12 are suicide cases and bars 13-22 are control samples.

"chr2_229931901_229954572_FBXO36_Validation.pdf". Taqman visualization from CopyCaller. Bars 1-7 are suicide cases and bars 8-17 are control samples.

"chr10_96521587_96521862_TM9SF3_Validation.tif". Gel visualization from PCR. Lane 1 ladder, lane 2-11 are suicide samples, lane 12 is a control sample, lane 13 ladder.

"chr12_25494302_25501878_LMNTD1_Validation.pdf". Taqman visualization from CopyCaller. Bars 1-19 are suicide cases and bars 20-29 are control samples.

"chr16_27325116_27339881_IL4R_Validation.pdf". Taqman visualization from CopyCaller. Bars 1-17 are suicide cases and bars 18-27 are control samples.

"chr16_83162541_83176121_CDH13_Validation.pdf". Taqman visualization from CopyCaller. Bars 1-20 are suicide cases and bars 21-36 are control samples.

"chr17_29056850_29058075_PIPOX_Validation.tif". Gel visualization from PCR. Lane 1 ladder, lane 2-9 are suicide samples, lane 10 is a control sample, lane 11 ladder.

"chr19_12292302_12294048_ZNF44_Validation.tif". Gel visualization from PCR. Lane 1 ladder, lane 2-16 are suicide samples, lane 17 is a control sample, lane 18 ladder.

"chr20_41184281_41184388_ZHX3_Validation.jpeg". Gel visualization from PCR. Lane 1 ladder, lane 2-7 are suicide samples, lane 8 is a control sample, lane 9 ladder.

"chr20_62294458_62295274_OSBPL2_Validation.jpeg" Gel visualization from PCR. Lane 1-2 ladder, lane 3-11 are suicide samples, lane 12 is a control sample, lane 13 ladder.

"chr22_37019285_37024652_MPST_Validation.tif". Gel visualization from PCR. Lane 1 ladder, lane 2-9 are suicide samples, lane 10 is a control sample, lane 11 ladder.

**GWAS Catalog References for each deletion**
**MPST**.
1. Chen MH, Raffield LM, Mousas A, Sakaue S, Huffman JE, Moscati A, Trivedi B, et al., Trans-ethnic and Ancestry-Specific Blood-Cell Genetics in 746,667 Individuals from 5 Global Populations. 2020, Cell.
2. Kurilshikov A , Medina-Gomez C , Bacigalupe R, Radjabzadeh D, Wang J, Demirkan A, Le Roy CI, et al., Large-scale association analyses identify host factors influencing human gut microbiome composition. 2021, Nat Genet.

3. Hysi PG, Mangino M , Christofidou P, Falchi M , Karoly ED, Nihr Bioresource Investigators, Mohney RP, et al., Metabolome Genome-Wide Association Study Identifies 74 Novel Genomic Regions Influencing Plasma Metabolites Levels. 2022, Metabolites.
4. Chen Y, Lu T, Pettersson-Kymmer U , Stewart ID, Butler-Laporte G, Nakanishi T, Cerani A, et al., Genomic atlas of the plasma metabolome prioritizes metabolites implicated in human diseases. 2023, Nat. Genet.
5. Hodonsky CJ , Jain D , Schick UM, Morrison JV, Brown L, McHugh CP , Schurmann C, Chen DD, et al., Genome-wide association study of red blood cell traits in Hispanics/Latinos: The Hispanic Community Health Study/Study of Latinos. 2017, PloS Genet.
6. Smith SM, Douaud G , Chen W , Hanayik T, Alfaro-Almagro F, Sharp K, Elliott LT. An expanded set of genome-wide association studies of brain imaging phenotypes in UK Biobank. 2021, Nat. Neurosci.
7. Sinnott-Armstrong N , Tanigawa Y, Amar D , Mars N, Benner C, Aguirre M, Venkataraman GR, et al., Genetics of 35 blood and urine biomarkers in the UK Biobank. 2021, Nat. Genet.

**<u>PIPOX</u>**.

1. Meng X, Navoly G, Giannakopoulou O, Levey DF, Koller D, Pathak GA, Koen N, et al., Multi-ancestry genome-wide association study of major depression aids locus discovery, fine mapping, gene prioritization and causal inference. 2024, Nat Genet.
2. Howard DM, Adams MJ, Clarke TK, Hafferty JD, Gibson J, Shirali M, Coleman JRI, et al., Genome-wide meta-analysis of depression identifies 102 independent variants and highlights the importance of the prefrontal brain regions. 2019, Nat. Neurosci.
3. Levey DF , Stein MB, Wendt FR, Pathak GA, Zhou H, Aslan M, Quaden R, et al., Bi-ancestral depression GWAS in the Million Veteran Program and meta-analysis in >1.2 million individuals highlight new therapeutic directions. 2021, Nat Neurosci.
4. Als TD, Kurki MI, Grove J , Voloudakis G, Therrien K , Tasanko E, Nielsen TT, Naamanka J, et al., Depression pathophysiology, risk prediction of recurrence and comorbid psychiatric disorders using genome-wide analyses. 2023, Nat Med.
5. Mitchell BL, Campos AI, Whiteman DC , Olsen CM, Gordon SD, Walker AJ, Dean OM, et al., The Australian Genetics of Depression Study: New Risk Loci and Dissecting Heterogeneity Between Subtypes. 2021, Biol Psychiatry.
6. Hyde CL, Nagle MW, Tian C, Chen X, Paciga SA, Wendland JR , Tung JY, et al., Identification of 15 genetic loci associated with risk of major depression in individuals of European descent. 2016, Nat. Genet.
7. Wang H, Yang J, Schneider JA, De Jager PL, Bennett DA, Zhang HY. Genome-wide interaction analysis of pathological hallmarks in Alzheimer's disease. 2020, Neurobiol Aging.
8. Christakoudi S, Evangelou E , Riboli E , Tsilidis KK. GWAS of allometric body-shape indices in UK Biobank identifies loci suggesting associations with morphogenesis, organogenesis, adrenal cell renewal and cancer. 2021. Sci Rep.
9. McCartney DL, Min JL, Richmond RC, Lu AT, Sobczyk MK, Davies G, Broer L, Genome-wide association studies identify 137 genetic loci for DNA methylation biomarkers of aging. 2021, Genome Biol.
10. Mekki Y, Guillemot V, Lemaitre H, Carrion-Castillo A , Forkel S, Frouin V , Philippe C, et al., The genetic architecture of language functional connectivity. 2021, Neuroimage.
11. Liu M, Jiang Y, Wedow R , Li Y , Brazel DM, Chen F, Datta G, et al., Association studies of up to 1.2 million individuals yield new insights into the genetic etiology of tobacco and alcohol use. 2019, Nat Genet.
12. Saunders GRB, Wang X, Chen F, Jang SK, Liu M , Wang C, Gao S, et al., Genetic diversity fuels gene discovery for tobacco and alcohol use. 2022, Nature.

**<u>FBXO36.</u>**

1. Rhee EP, Ho JE , Chen MH, Shen D, Cheng S, Larson MG , Ghorbani A, et al., A genome-wide association study of the human metabolome in a community-based cohort. 2013, Cell Metabol.
2. Sakaue S, Kanai M , Tanigawa Y, Karjalainen J, Kurki M, Koshiba S, Narita A, et al. A cross-population atlas of genetic associations for 220 human phenotypes. 2021, Nat. Genet.
3. Harris BHL, Di Giovannantonio M, Zhang P, Harris DA, Lord SR, Allen NE, Maughan TS, et al. New role of fat-free mass in cancer risk linked with genetic predisposition. 2024, Sci. Rep.

4. Hawkes G, Beaumont RN, Tyrrell J, Power GM, Wood A, Laakso M, Silva LF, et al. Genetic evidence that high BMI in childhood has a protective effect on intermediate diabetes traits, including measures of insulin sensitivity and secretion, after accounting for BMI in adulthood. 2023, Diabetologia

**LMNTD1.**
1. Rühlemann MC , Hermes BM, Bang C, Doms S , Moitinho-Silva L, Thingholm LB, Frost F, et al. Genome-wide association study in 8,956 German individuals identifies influence of ABO histo-blood groups on gut microbiome. 2021, Nat. Genet.
2. Sakaue S, Kanai M , Tanigawa Y, Karjalainen J, Kurki M, Koshiba S, Narita A, et al. A cross-population atlas of genetic associations for 220 human phenotypes. 2021, Nat. Genet.

**TM9SF3.**
1. Yao Y, Chu X, Ma M, Ye J, Wen Y, Li P, Cheng B, et al. Evaluate the effects of serum urate level on bone mineral density: a genome-wide gene-environment interaction analysis in UK Biobank cohort. 2021, Endocrine.

**ZNF44.**
1. Kim KY , Kim JO, Kim YS, Choi JE, Park JM, Han K, Park DH, et al. Genome-wide association of individual vulnerability with alcoholic liver disease: A Korean Genome and Epidemiology Study. 2021, Hepatology.

**OSBPL2.**
1. Mullins N, Forstner AJ, O'Connell KS , Coombes B, Coleman JRI, Qiao Z, Als TD, et al., Genome-wide association study of more than 40,000 bipolar disorder cases provides new insights into the underlying biology. 2021, Nat. Genet.
2. Chen MH, Raffield LM, Mousas A, Sakaue S, Huffman JE, Moscati A, Trivedi B, et al. Trans-ethnic and Ancestry-Specific Blood-Cell Genetics in 746,667 Individuals from 5 Global Populations. 2020, Cell.
3. Gelernter J , Kranzler HR, Sherva R, Almasy L, Herman AI, Koesterer R, Zhao H, et al. Genome-wide association study of nicotine dependence in American populations: identification of novel risk loci in both African-Americans and European-Americans. 2014, Biol Psychiatry

**ZHX3.**
https://www.ebi.ac.uk/gwas/genes/ZHX3

**CLCA4.**
https://www.ebi.ac.uk/gwas/genes/CLCA4

**IL4R.**
https://www.ebi.ac.uk/gwas/genes/IL4R

**CDH13.**
https://www.ebi.ac.uk/gwas/genes/CDH13
