## Supplementary figures and images for "Intragenic deletions from whole genome sequencing of 1054 suicide deaths"

### chr1_86562530_86573603_CLCA4_Samplot.png

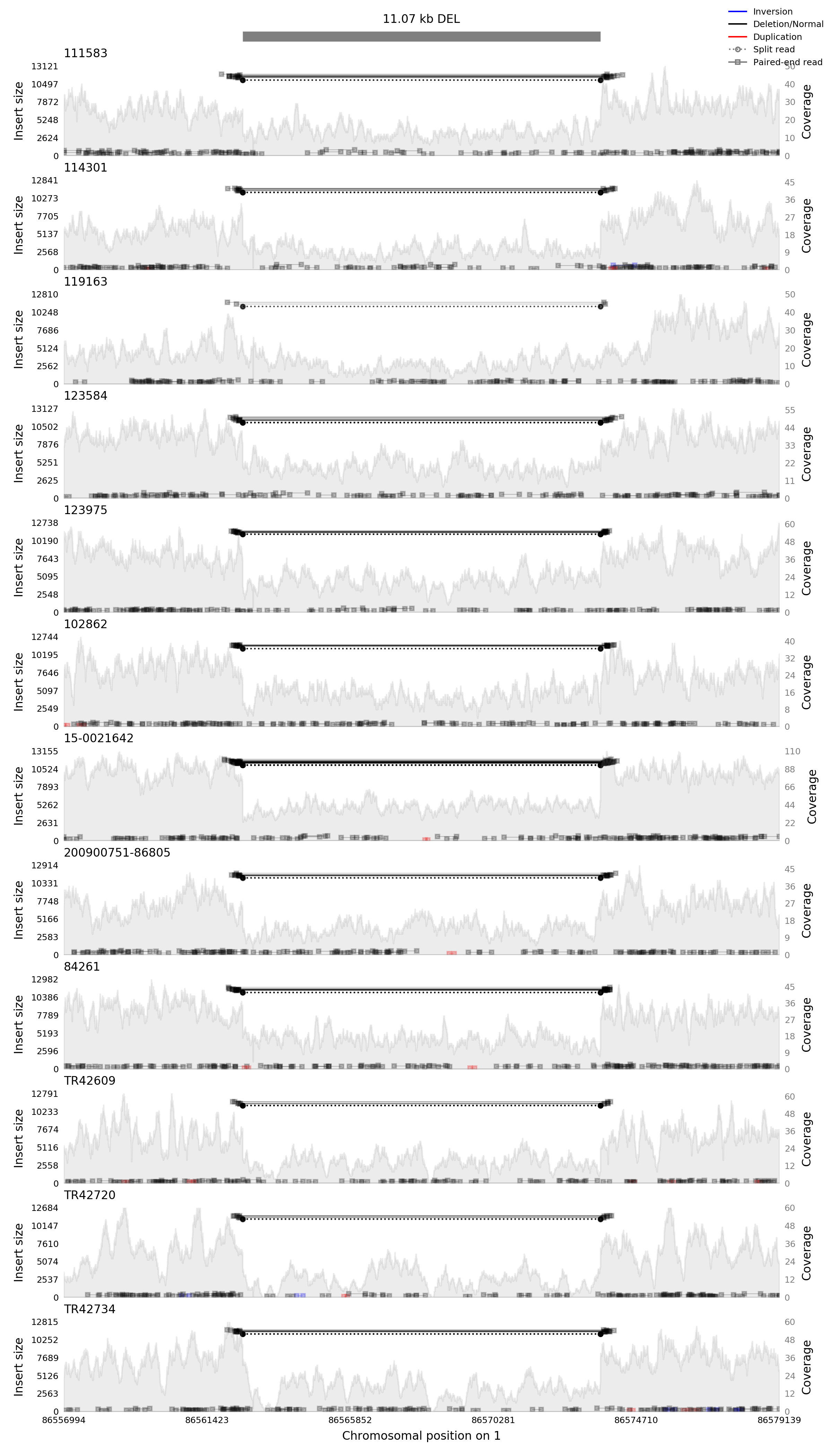

### chr1_86562530_86573603_CLCA4_Validation.pdf

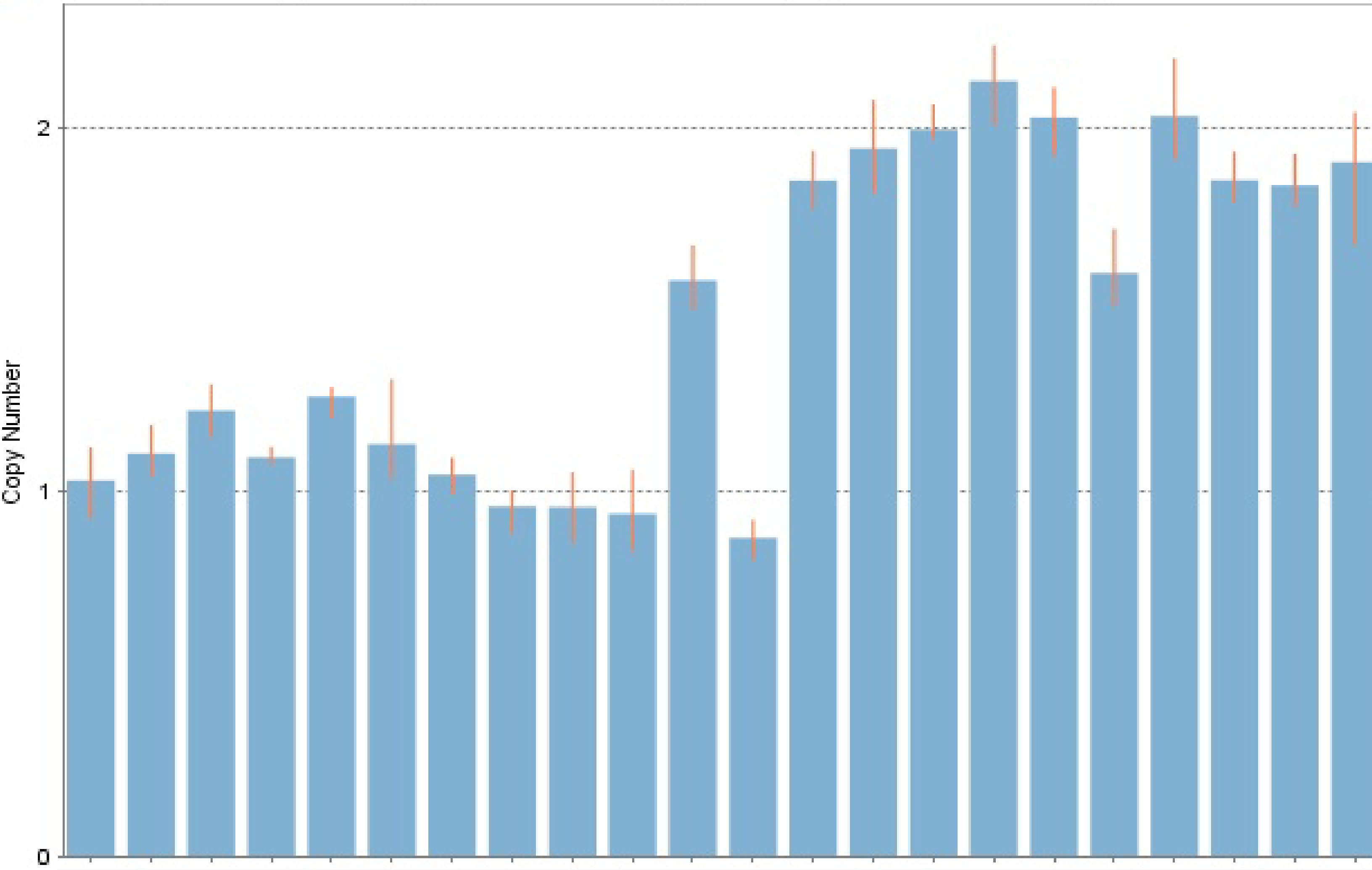

### chr2_229931901_229954572_FBXO36_Samplot.png

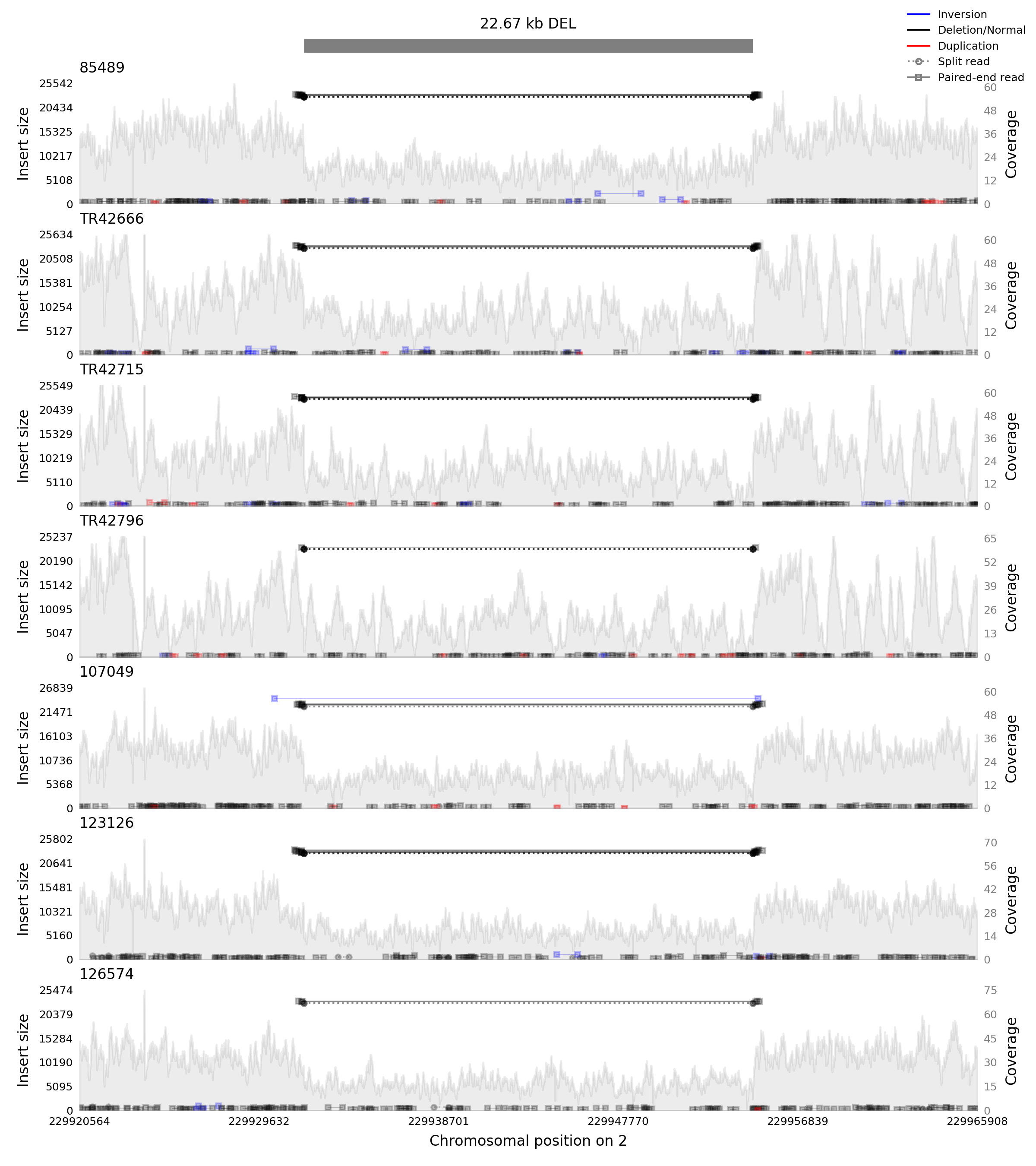

### chr2_229931901_229954572_FBXO36_Validation.pdf

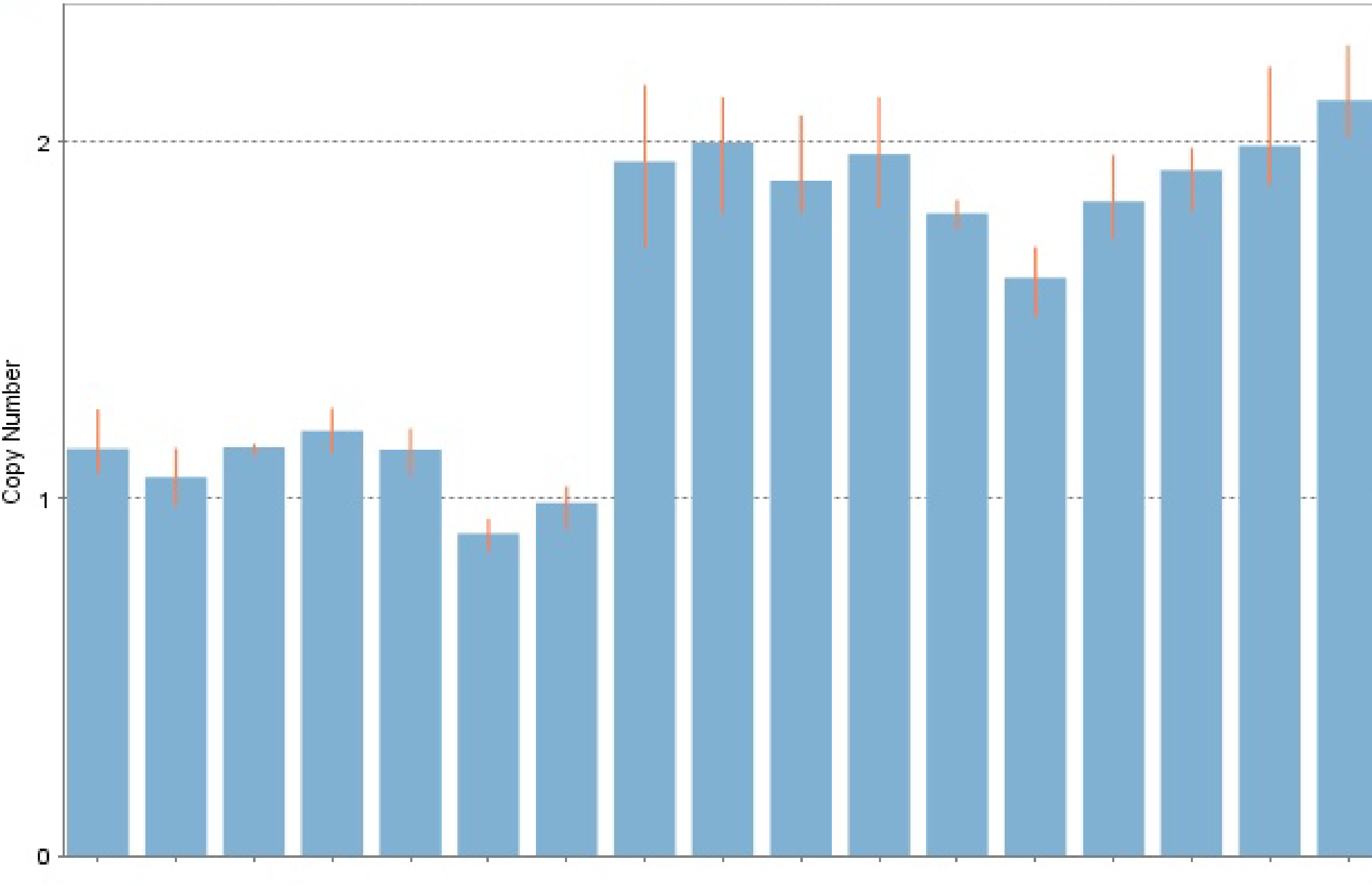

### chr10_96521587_96521862_TM9SF3_Samplot.png

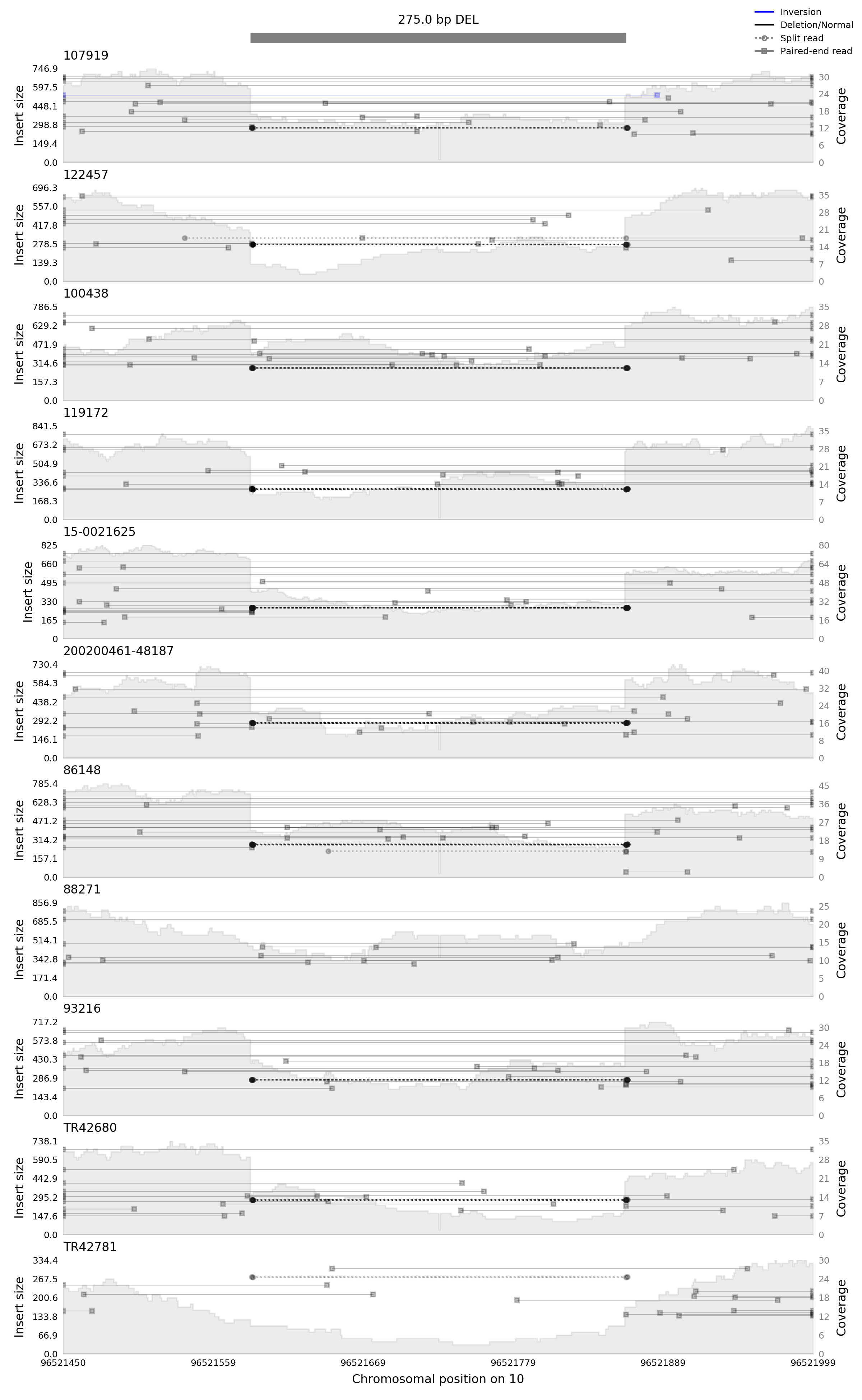

### chr10_96521587_96521862_TM9SF3_Validation.tif

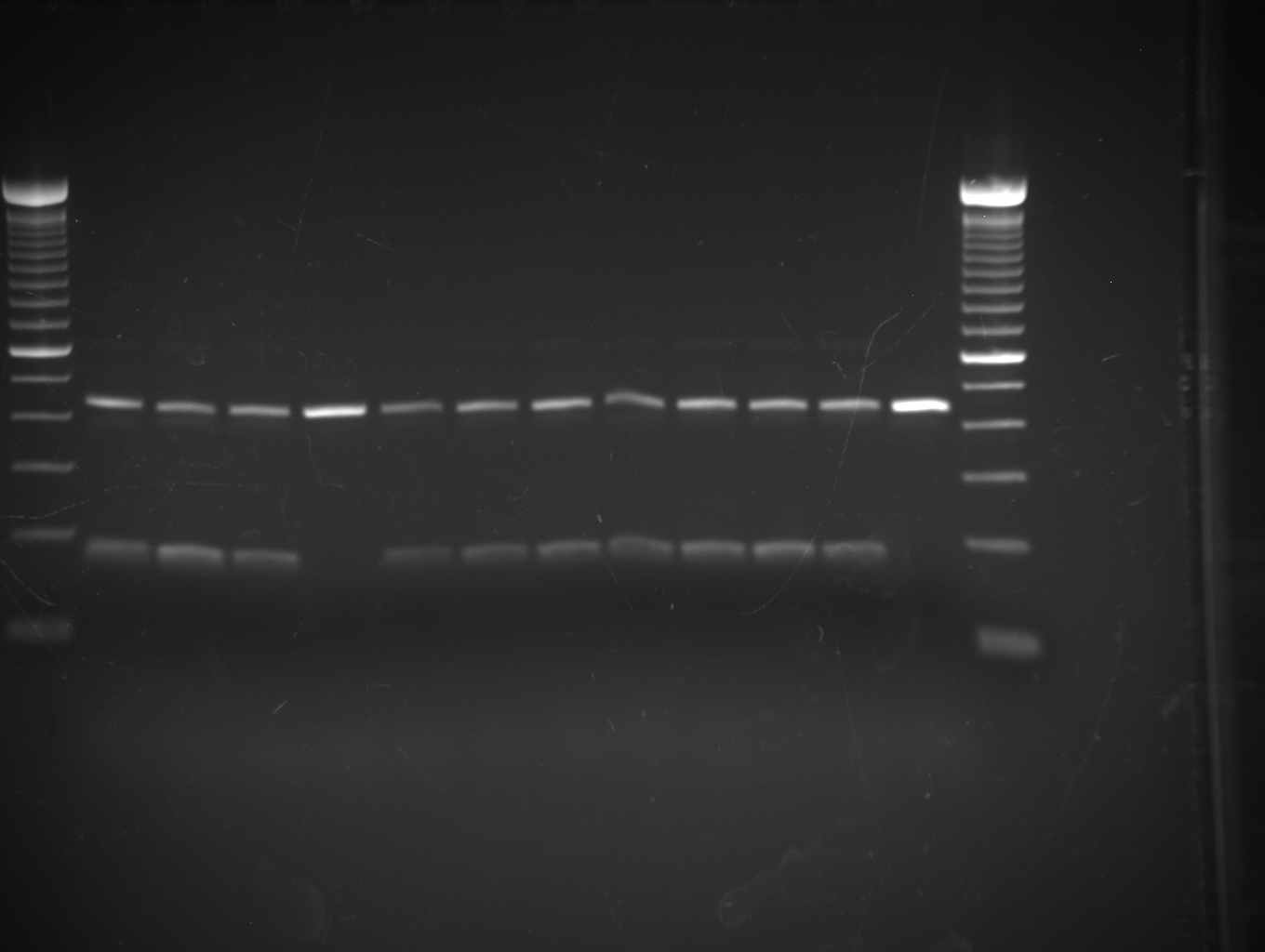

### chr12_25494302_25501878_LMNTD1_Samplot.png

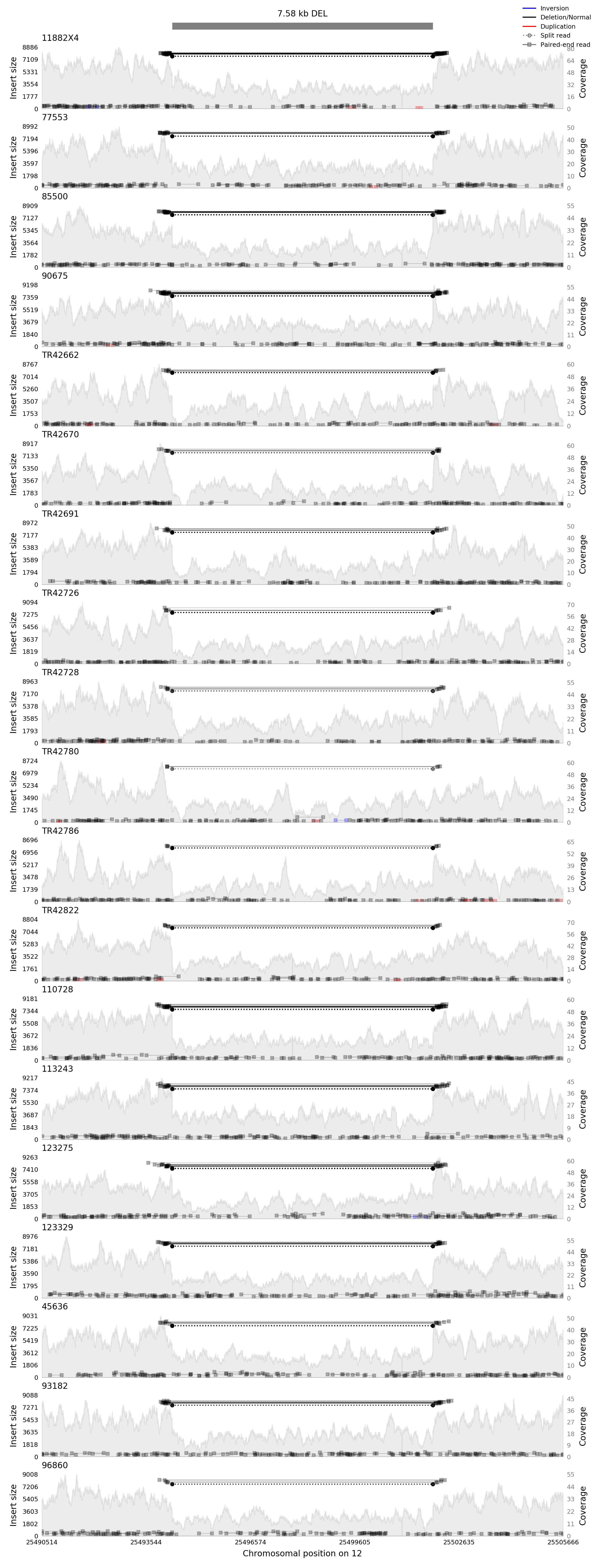

### chr12_25494302_25501878_LMNTD1_Validation.pdf

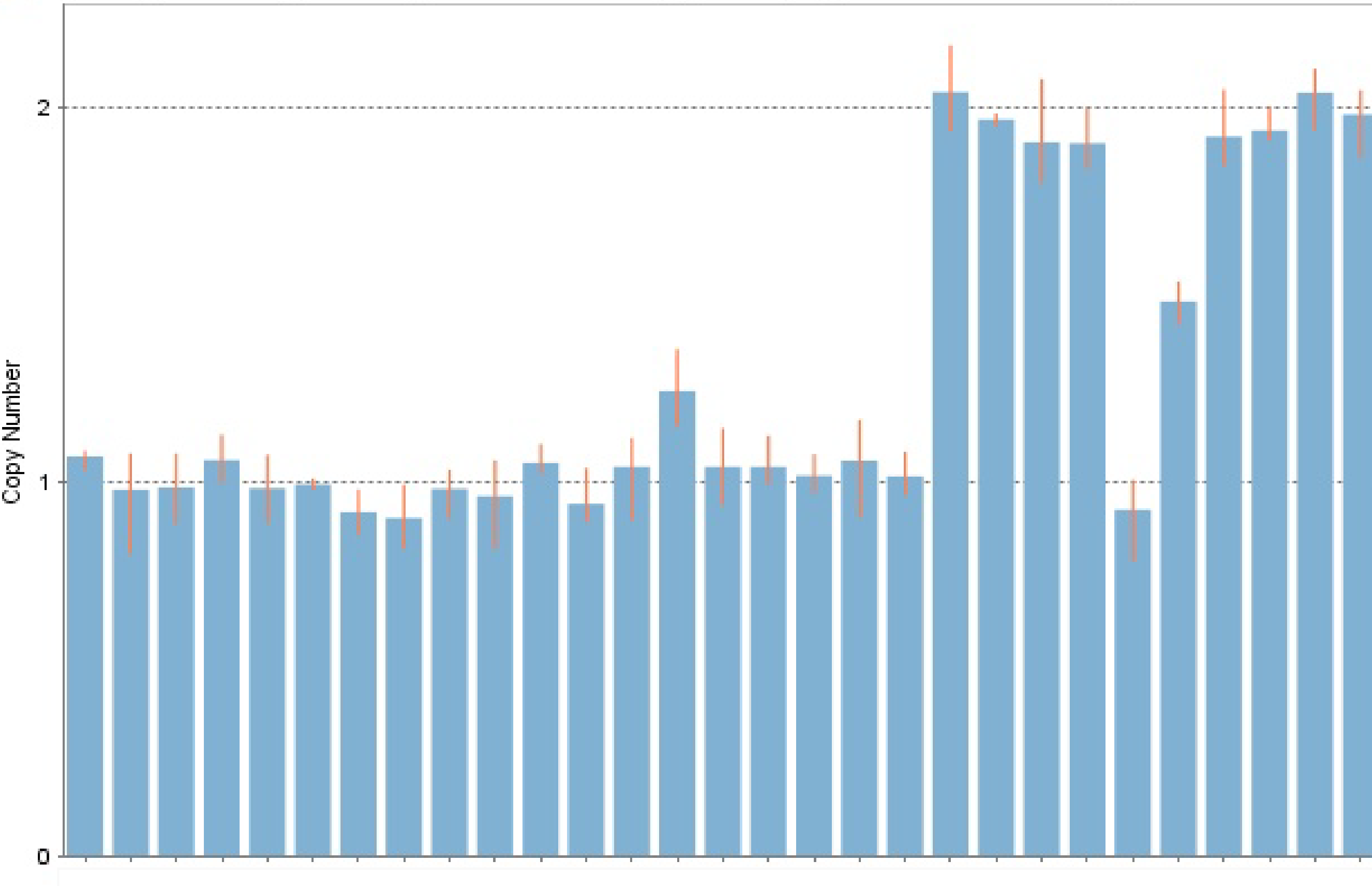

### chr16_27325116_27339881_IL4R_Samplot.png

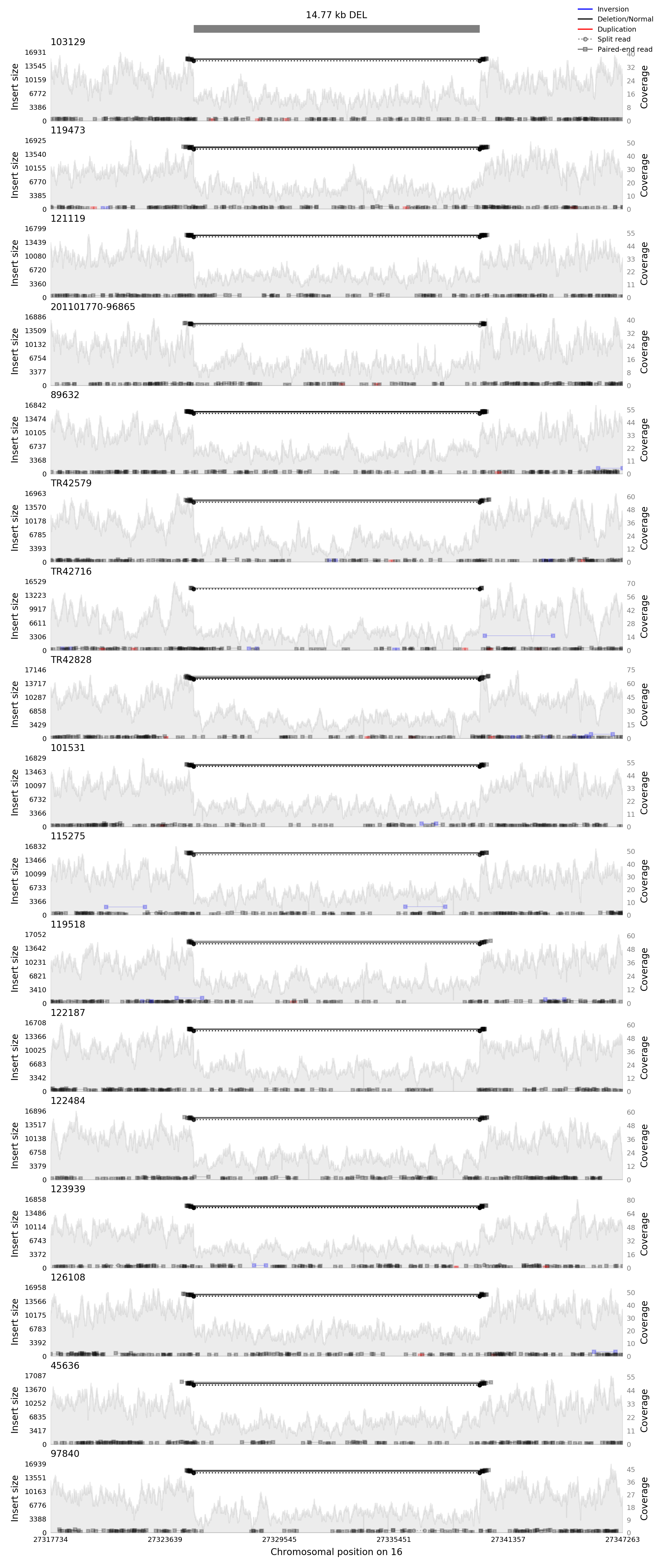

### chr16_27325116_27339881_IL4R_Validation.pdf

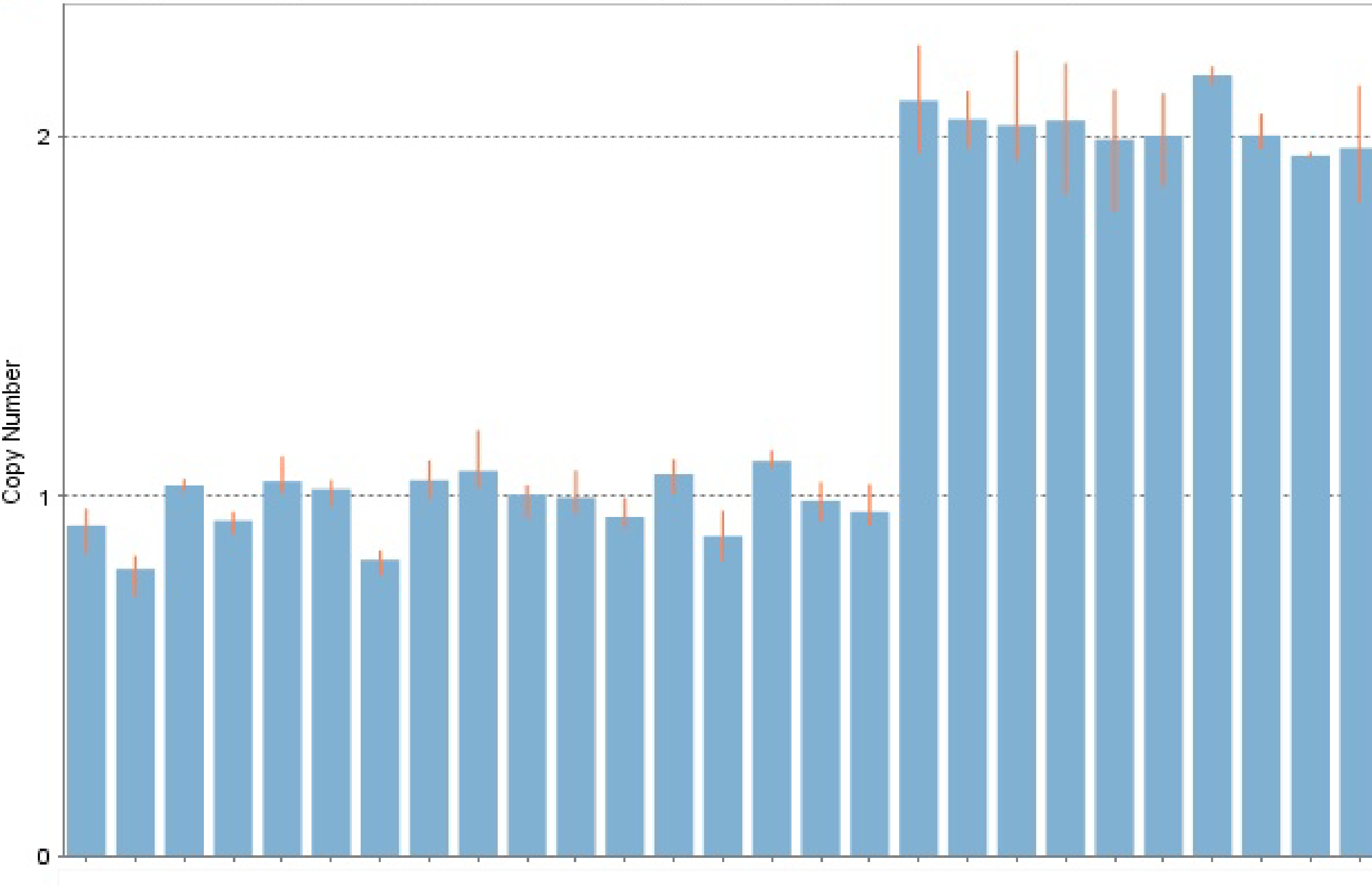

### chr16_83162541_83176121_CDH13_Samplot.png

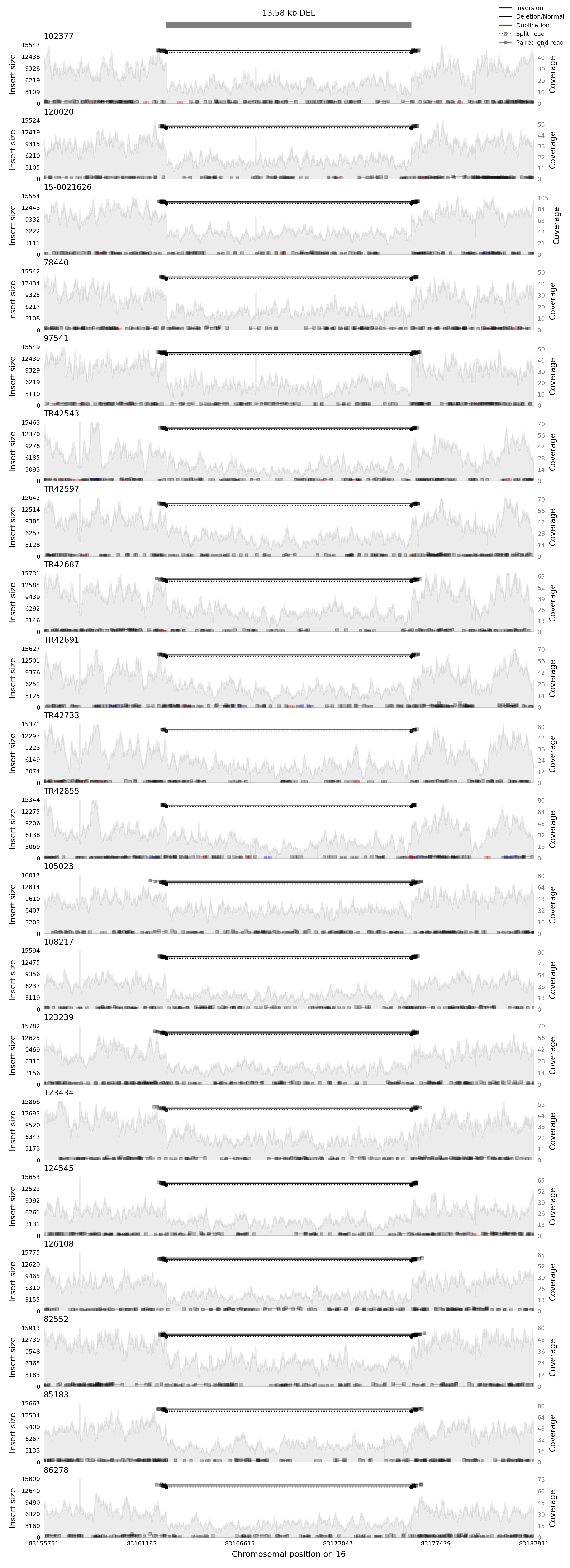

### chr16_83162541_83176121_CDH13_Validation.pdf

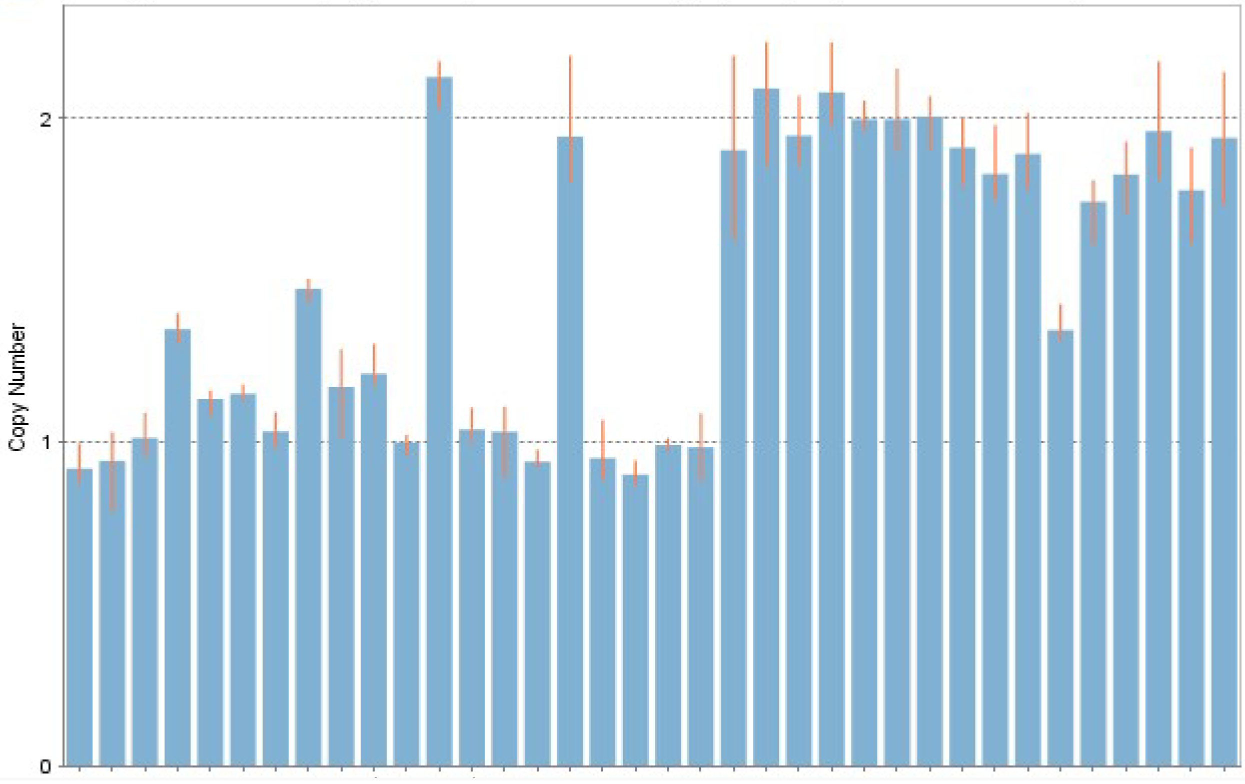

### chr17_29056850_29058075_PIPOX_Samplot.png

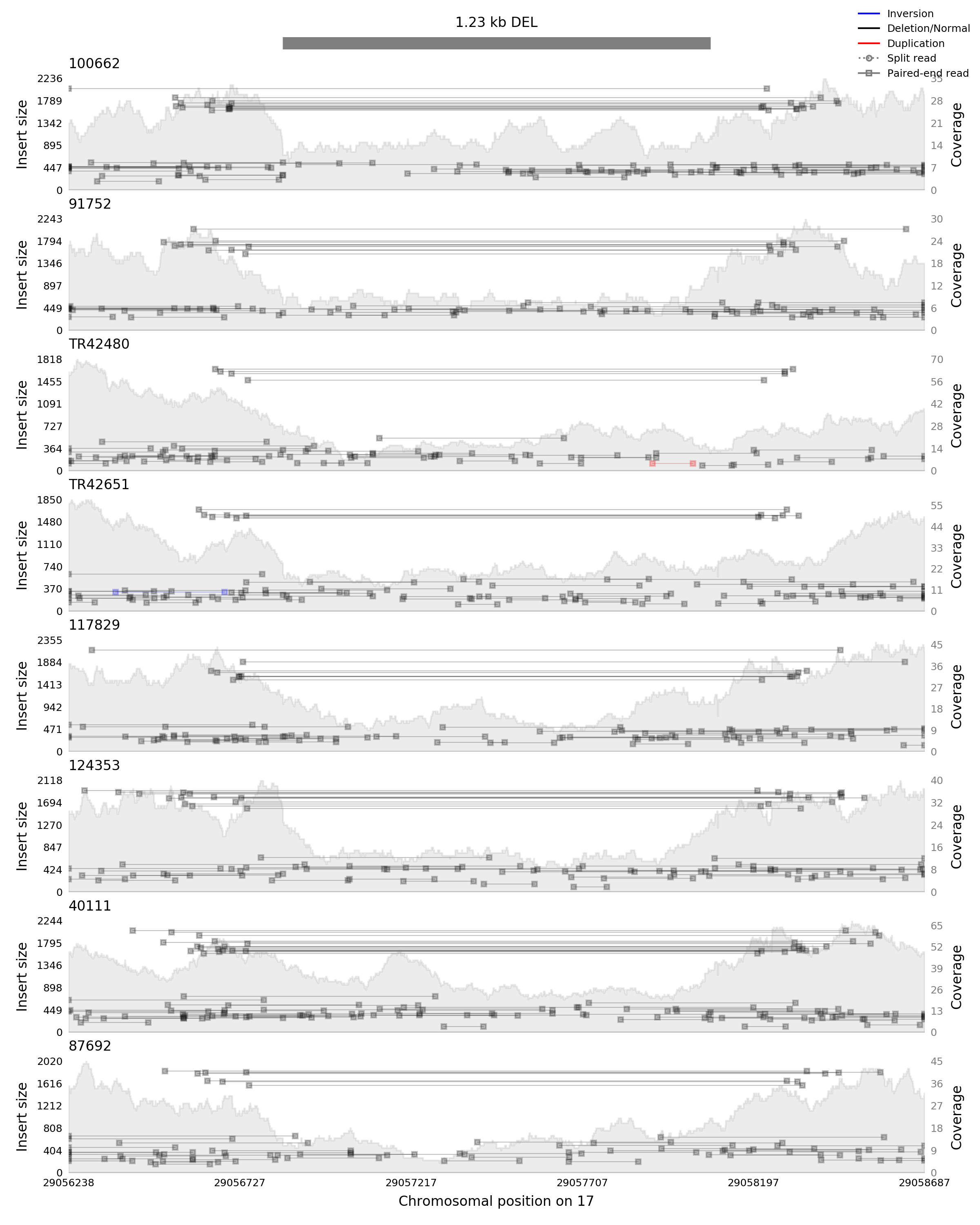

### chr17_29056850_29058075_PIPOX_Validation.tif

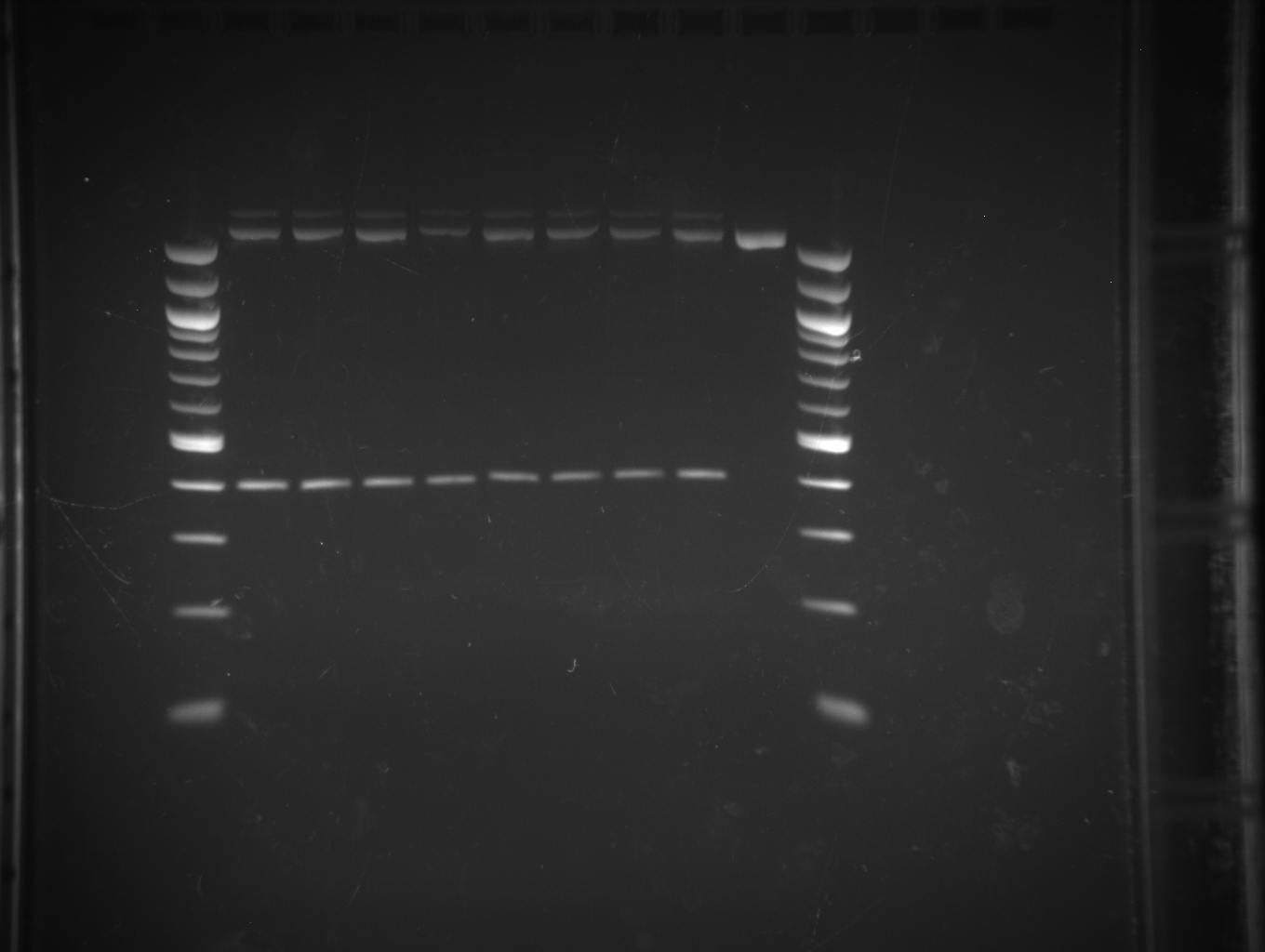

### chr19_12292302_12294048_ZNF44_Samplot.png

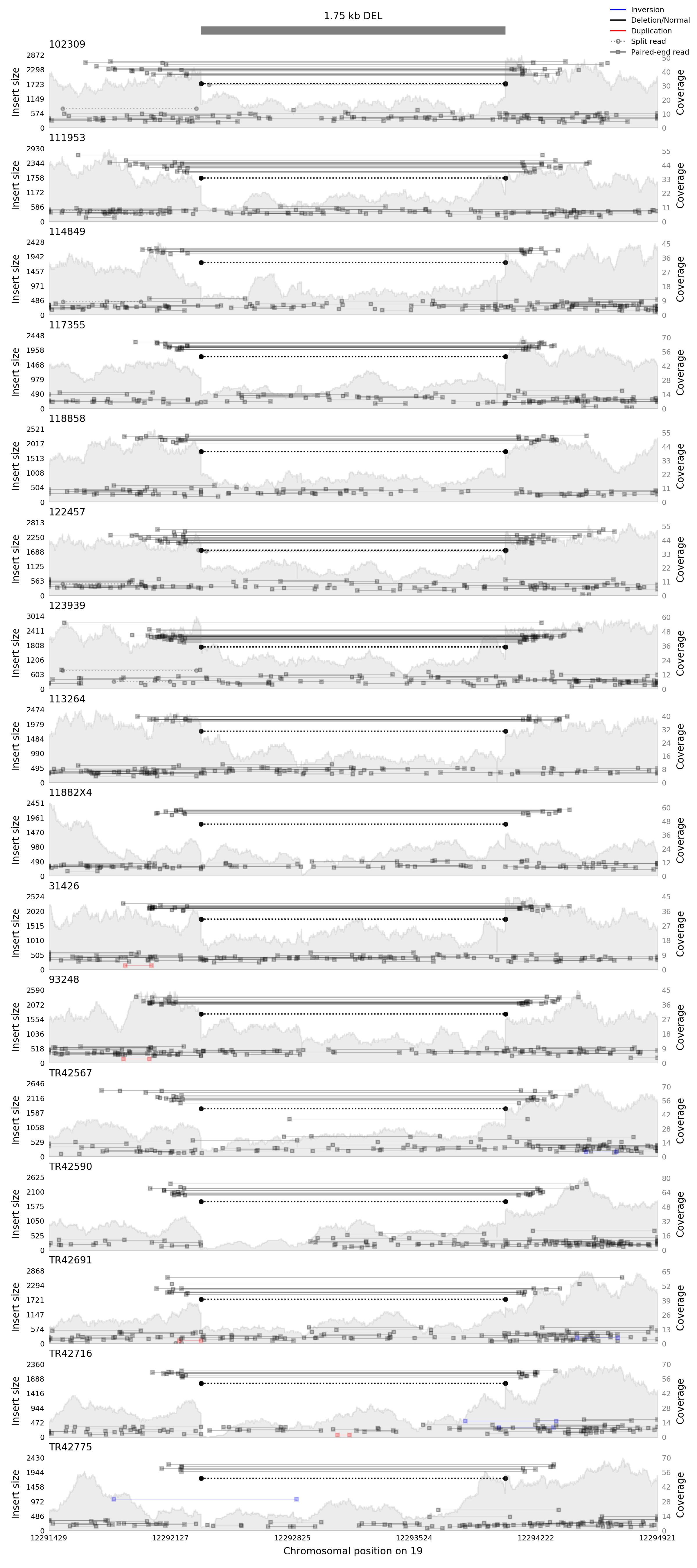

### chr19_12292302_12294048_ZNF44_Validation.tif

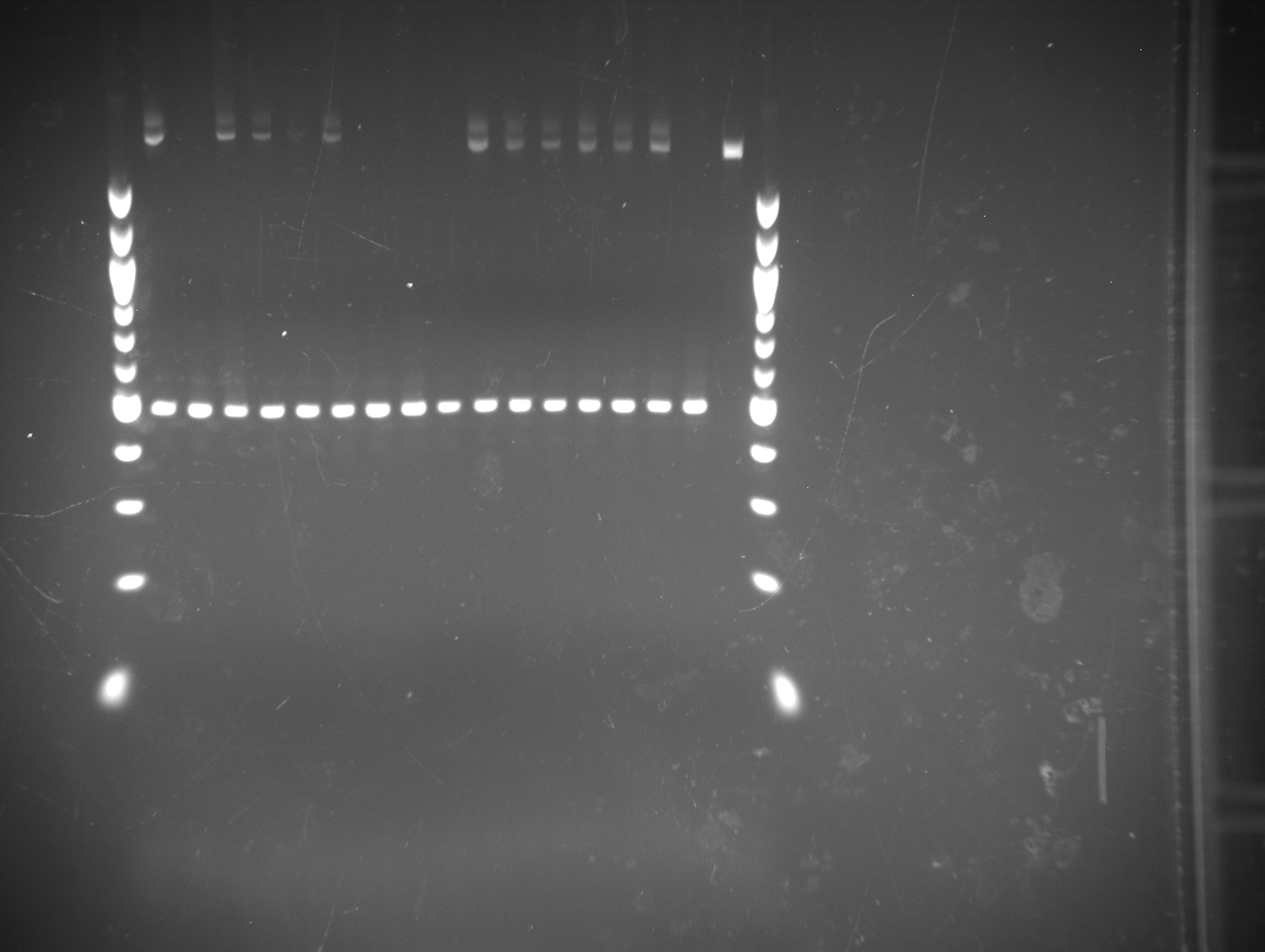

### chr20_41184281_41184388_ZHX3_Samplot.png

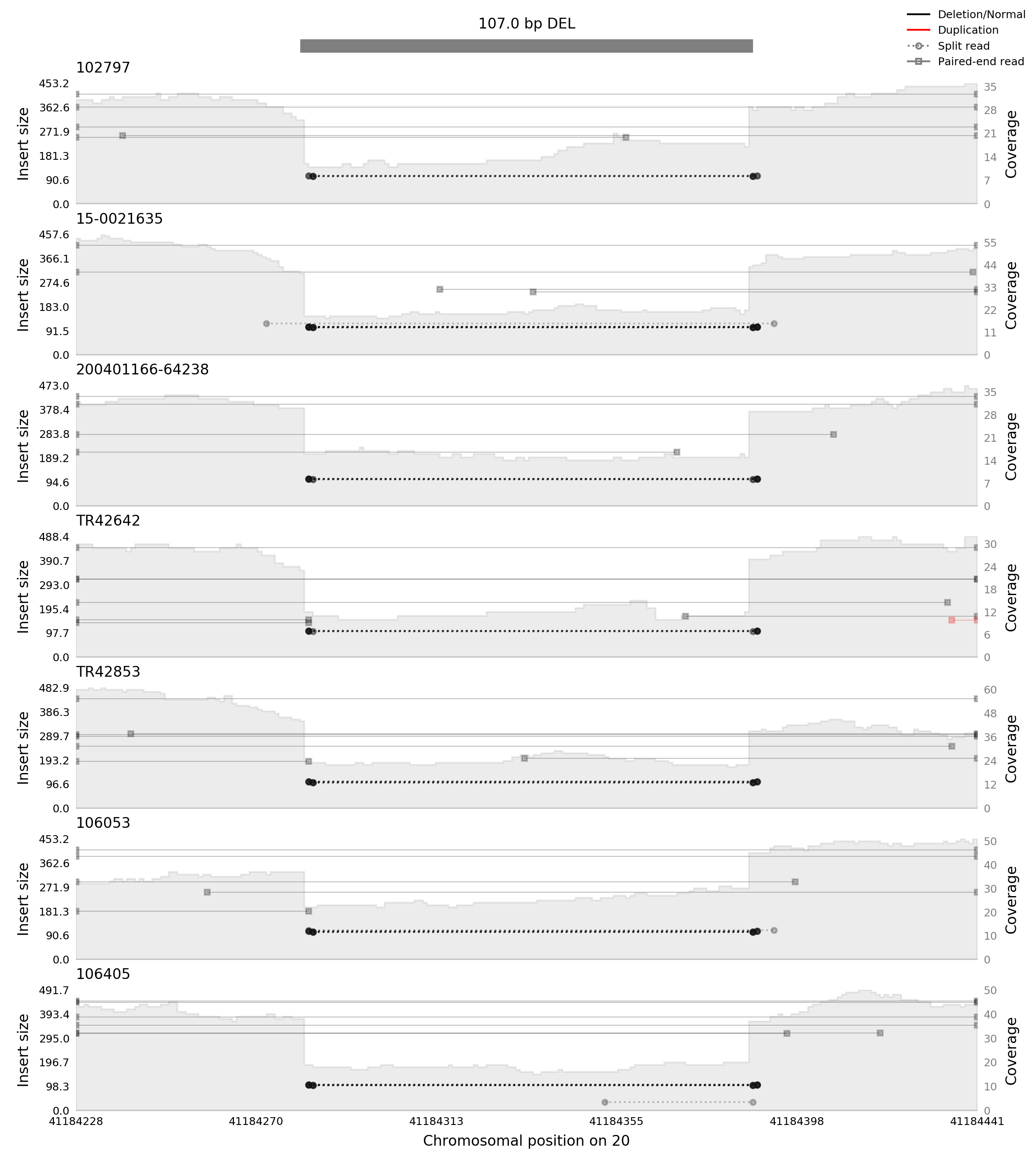

### chr20_41184281_41184388_ZHX3_Validation.jpeg

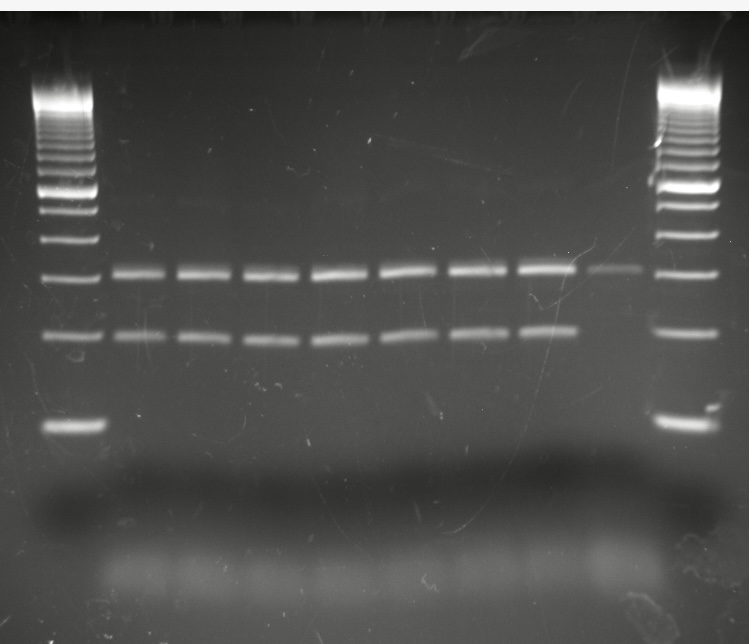

### chr20_62294458_62295274_OSBPL2_Samplot.png

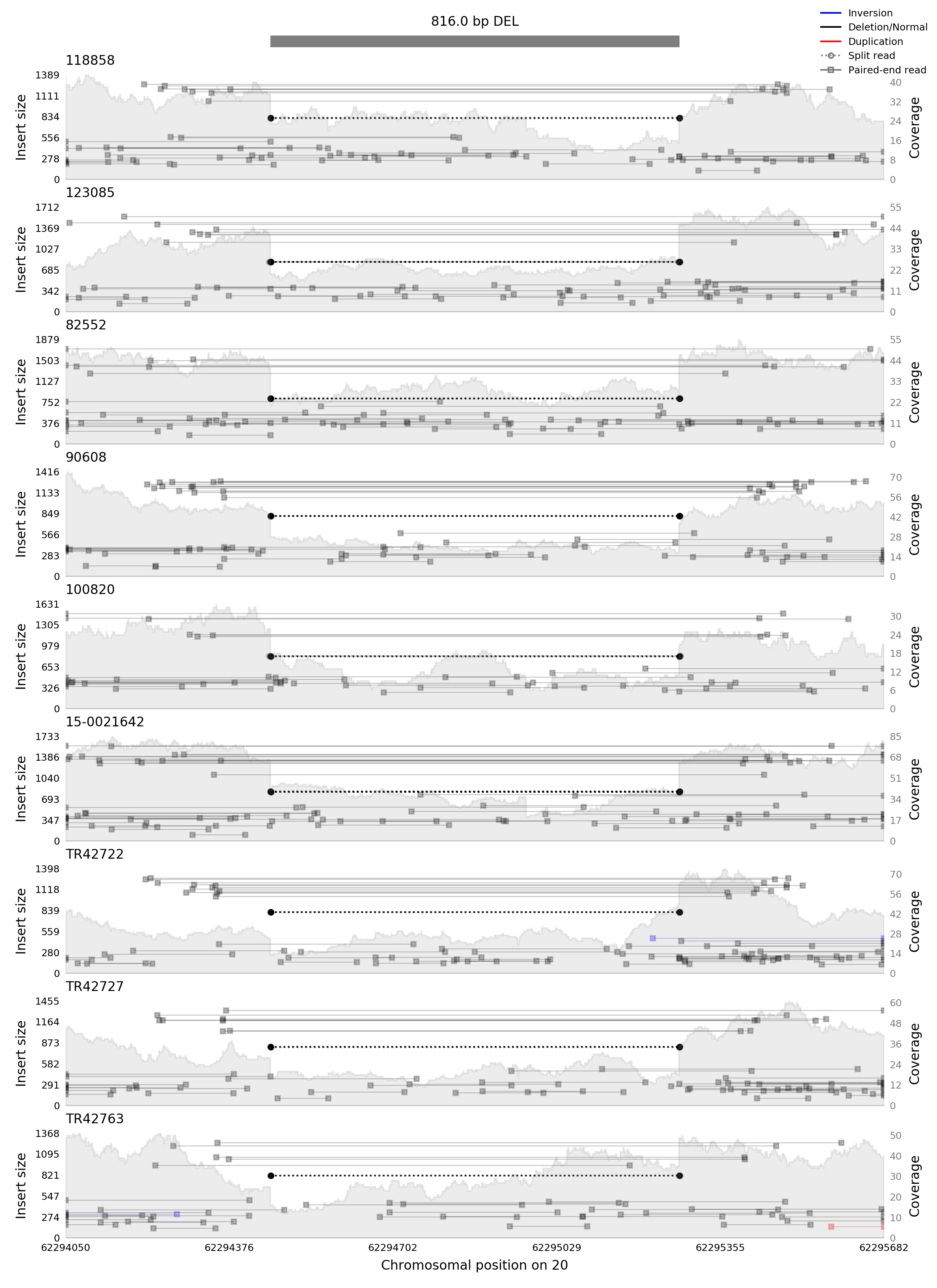

### chr20_62294458_62295274_OSBPL2_Validation.jpeg

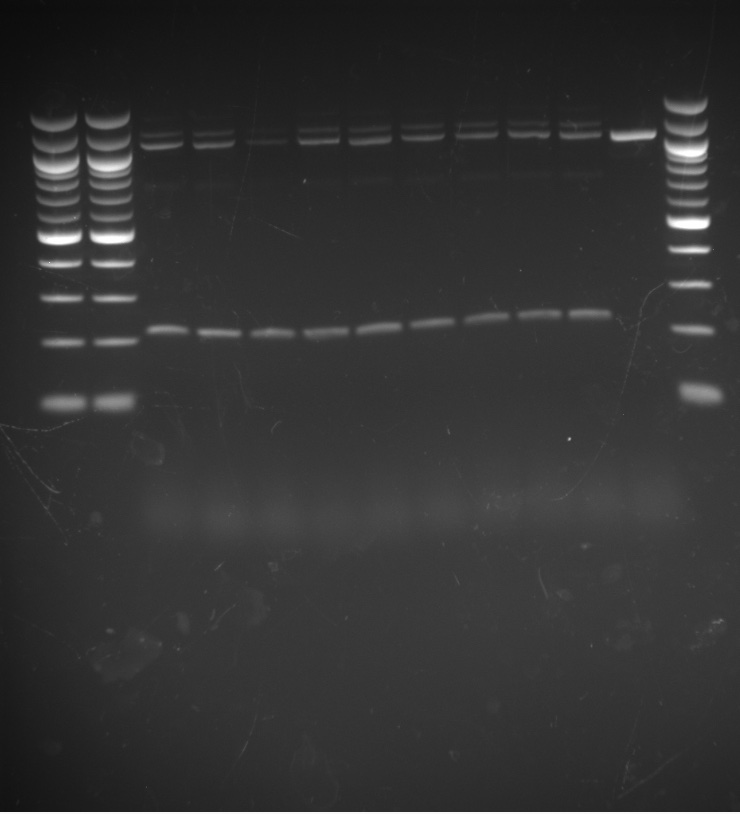

### chr22_37019285_37024652_MPST_Samplot.png

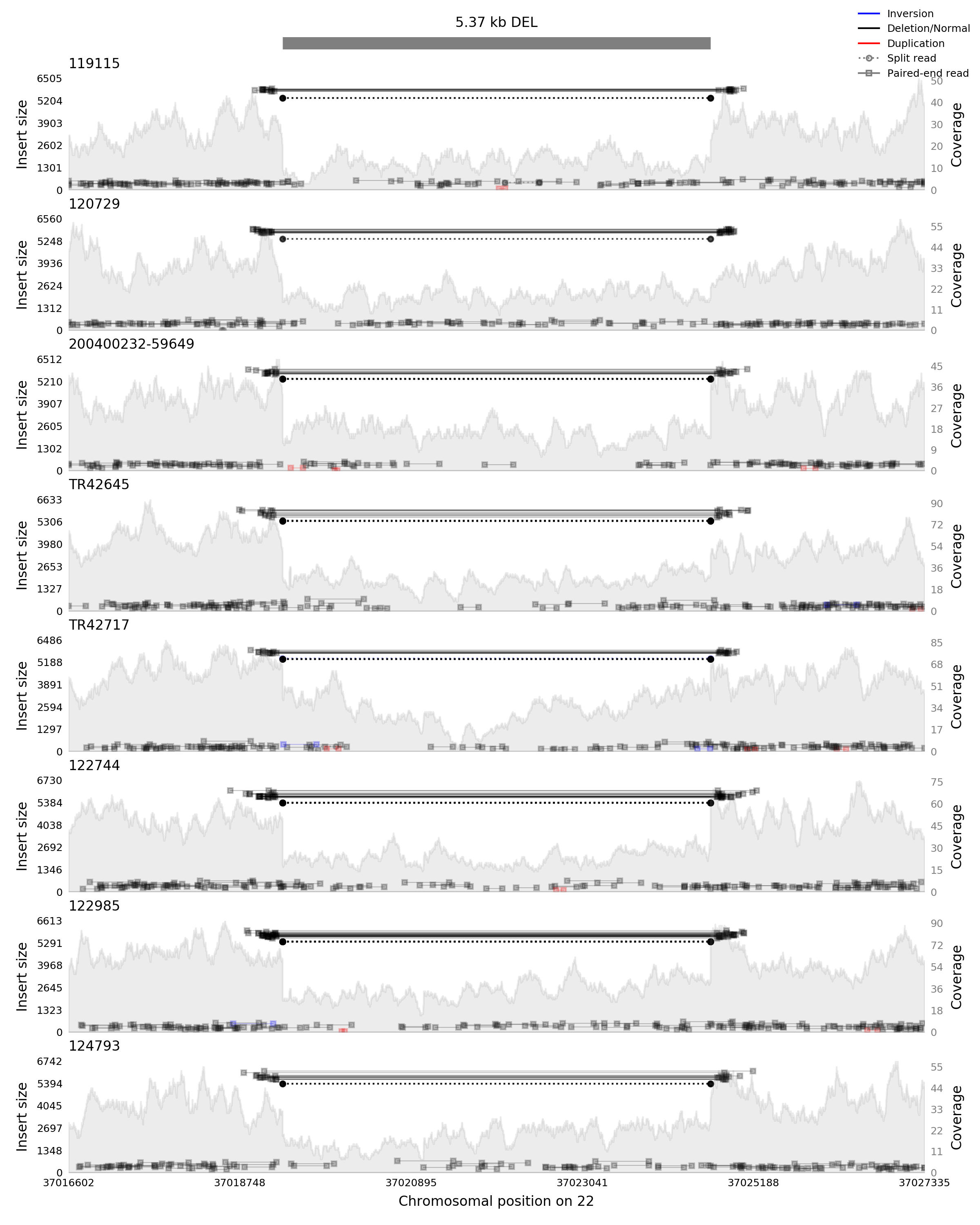

### chr22_37019285_37024652_MPST_Validation.tif

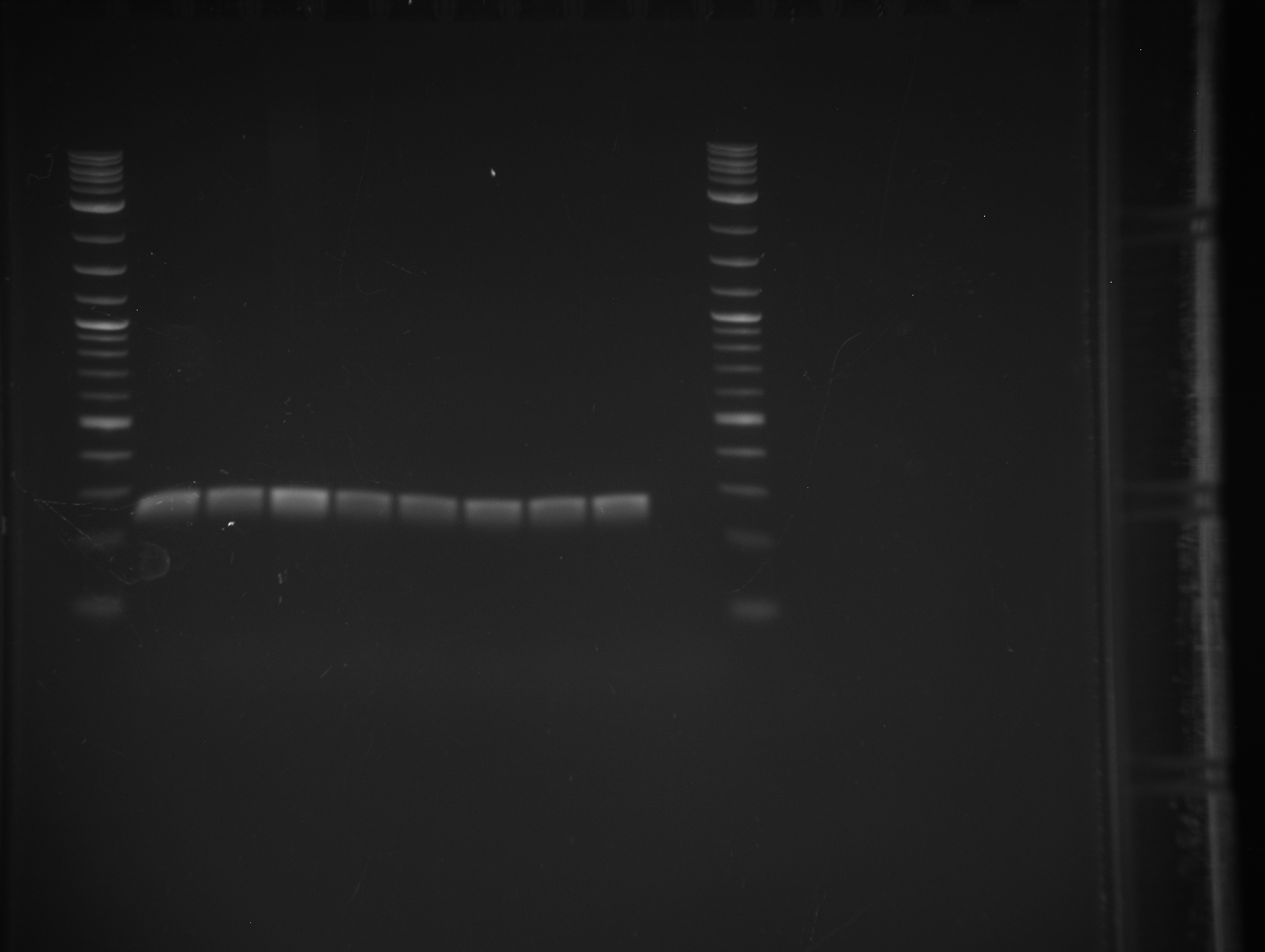
